# Blood biomarkers representing maternal-fetal interface tissues used to predict early-and late-onset preeclampsia but not COVID-19 infection

**DOI:** 10.1101/2022.06.09.22276209

**Authors:** Herdiantri Sufriyana, Hotimah Masdan Salim, Akbar Reza Muhammad, Yu-Wei Wu, Emily Chia-Yu Su

## Abstract

**Background:** A well-known blood biomarker (soluble fms-like tyrosinase-1 [sFLT-1]) for preeclampsia, i.e., a pregnancy disorder, was found to predict severe COVID-19, including in males. True biomarker may be masked by more-abrupt changes related to endothelial instead of placental dysfunction. This study aimed to identify blood biomarkers that represent maternal-fetal interface tissues for predicting preeclampsia but not COVID-19 infection.

**Methods:** The surrogate transcriptome of the tissues was determined by that in maternal blood, utilizing four datasets (*n*=1,354) which were collected before the COVID-19 pandemic. Applying machine learning, a preeclampsia prediction model was chosen between those using blood transcriptome (differentially expressed genes [DEGs]) and the blood-derived surrogate for the tissues. We selected the most predictive model by the area under receiver operating characteristic (AUROC) using a dataset for developing the model, and well-replicated in datasets either with or without intervention. To identify eligible blood biomarkers that predicted any-onset preeclampsia from the datasets but did not predict positives in the COVID-19 dataset (*n*=47), we compared several methods of predictor discovery: (1) the best prediction model; (2) gene sets by standard pipelines; and (3) a validated gene set for predicting any-onset preeclampsia during the pandemic (*n*=404). We chose the most predictive biomarkers from the best method with the significantly largest number of discoveries by a permutation test. The biological relevance was justified by exploring and reanalyzing low- and high-level, multi-omics information.

**Results:** A prediction model using the surrogates developed for predicting any-onset preeclampsia (AUROC of 0.85, 95% confidence interval [CI] 0.77 to 0.93) was the only that was well-replicated in an independent dataset with no intervention. No model was well-replicated in datasets with a vitamin D intervention. None of the blood biomarkers with high weights in the best model overlapped with blood DEGs. Blood biomarkers were transcripts of integrin-α5 (ITGA5), interferon regulatory factor-6 (IRF6), and P2X purinoreceptor-7 (P2RX7) from the prediction model, which was the only method that significantly discovered the eligible blood biomarkers (*n*=3/100 combinations, 3.0%; *P*=.036). Most of the predicted events (73.70%) among any-onset preeclampsia were cluster A as defined by ITGA5 (Z-score ≥1.1), but were only a minority (6.34%) among positives in the COVID-19 dataset. The remaining were the predicted events (26.30%) among any-onset preeclampsia or those among COVID-19 infection (93.66%) if IRF6 Z-score was ≥-0.73 (clusters B and C), in which none was the predicted events among either late-onset preeclampsia (LOPE) or COVID-19 infection if P2RX7 Z-score was <0.13 (cluster B). Greater proportion of predicted events among LOPE were cluster A (82.85% vs. 70.53%) compared to early-onset preeclampsia (EOPE). The biological relevance by multi-omics information explained the biomarker mechanism, polymicrobial infection in any-onset preeclampsia by ITGA5, viral co-infection in EOPE by ITGA5-IRF6, a shared prediction with COVID-19 infection by ITGA5-IRF6-P2RX7, and non-replicability in datasets with a vitamin D intervention by ITGA5.

**Conclusions:** In a model that predicts preeclampsia but not COVID-19 infection, the important predictors were maternal-blood genes that were not extremely expressed, including the proposed blood biomarkers. The predictive performance and biological relevance should be validated in future experiments.

## 1. Introduction

Preeclampsia is a two-stage disorder consisting of placental and endothelial dysfunction [1]. The latter is shared with other disorders and diseases, and is not limited to placental dysfunction-related diseases such as preeclampsia [2]. This may lead to false discovery of predictive biomarkers for preeclampsia particularly in terms of blood biomarkers. For instance, soluble fms-like tyrosinase-1 (sFlt-1) is a well-known predictor of early-onset preeclampsia (EOPE), especially during the first trimester of pregnancy [3]. However, since recent evidence also showed that sFlt-1 could predict severe cases of COVID-19 [4, 5], it is unclear whether sFlt-1 is specific to preeclampsia or any endothelial dysfunction-related diseases.

Hypertension in pregnancy, including preeclampsia (3%∼8% of pregnancies) [6], is an emerging cause of maternal deaths worldwide [7]. Although EOPE can be predicted and prevented, this subtype only contributes to ∼10% of cases of preeclampsia [8]. While it is less severe than EOPE, pregnant women with the late-onset subtype (late-onset preeclampsia [LOPE]) have doubled the risk compared to those without preeclampsia (adjusted odds ratio [aOR] 1.7, 95% confidence interval [CI] 1.6 to 1.9) in terms of severe maternal morbidity (5.5 vs. 3.0 per 100 deliveries) and mortality (11.2 vs. 4.2 per 100,000 deliveries) [9]. Late-onset, preterm preeclampsia also contributes to perinatal morbidity by medically induced prematurity, since the only cure is early delivery [1], particularly in ca. 70% of cases that are severe preeclampsia [10]. Working in tandem with a low-cost high-sensitivity prediction model [11], a specific prediction (i.e. with low false positives) is needed to avoid a false decision to delivery early, leading to medically induced prematurity. This is particularly true among babies from preeclamptic women and those with fetal growth restriction (FGR) from normotensive women, which share common predictors [12]. In addition to FGR, preeclampsia also shares a common pathogenesis with spontaneous preterm delivery, but both require opposite clinical interventions [13, 14]. The coronavirus disease 2019 (COVID-19) pandemic may also increase false positives [4, 5]. Therefore, finding blood biomarkers for any-onset preeclampsia is crucial to develop strategies for predicting and preventing preeclampsia in order to improve both maternal and perinatal outcomes of pregnancy, including ones that do not lead to false positives due to COVID-19 infection.

A review of 126 systematic reviews of preeclampsia predictions found that the most consistent blood biomarkers were placenta growth factor which was particularly relevant for first-trimester predictions of EOPE, and sFlt-1 which had a stronger association when tested later in the pregnancy [15]. The latter blood biomarker was also found to predict severe COVID-19 [4, 5], which implied that both preeclampsia and COVID-19 shared common mechanisms of endothelial dysfunction [16, 17]. To predict preeclampsia, particularly regardless of the onset [8], gene expression signatures of preeclampsia were widely studied in maternal-fetal interface tissues [18]. None of the transcriptomes identified in the tissues was included in blood protein biomarkers to predict preeclampsia [19]. Subtle changes in blood biomarkers may occur that correspond to changes in maternal-fetal interface tissues [20]. Nevertheless, these may be masked by more-abrupt changes related to endothelial instead of placental dysfunction, probably due to methodological limitations of differential expression analyses which reveal only extremely expressed genes. A recent study investigated early-pregnancy placental transcriptome signatures of preeclampsia, which led to global blood biomarkers unique for predicting this disease at any onset [21]. However, it is unclear whether the biomarkers significantly differ future preeclampsia from COVID-19 infection.

To identify clinically useful biomarkers that predict any-onset preeclampsia, several conditions should apply. We need a biomarker that can be sampled from the blood but represents a condition in maternal-fetal interface tissues. The surrogate transcriptome of those tissues, as inferred from the blood transcriptome, would subsequentially be utilized to develop a multivariable model that predict any-onset preeclampsia. This should be compared between independent cohorts with and without a particular early intervention; thus, potential preventive strategies can be proposed by an explanatory instead of exploratory approach to avoid confirmatory bias from investigators. A prediction model should be generalized in terms of both true positive and negative rates if it is replicated by an independent cohort with no intervention, but it might not be replicated in a cohort with a particular intervention. This is because the latter will likely have a different causal structure due to the intervention effect; however, the model should be unique to a positive outcome which is any-onset preeclampsia. Shared predictions with COVID-19 infection should be avoided, covering asymptomatic, and mild and severe symptomatic COVID-19, because this condition may co-exist with preeclampsia in any-trimester pregnant women; thus, this may lead to false positives of an early prediction of preeclampsia, especially in the presence of asymptomatic COVID-19. Eventually, only a few potential blood biomarkers should be inferred from the model to allow low-cost, practical implementation in clinical settings. This study aimed to identify potential blood biomarkers that represent the surrogate transcriptome of maternal-fetal interface tissues based on a model that predicts EOPE and LOPE but not COVID-19 infection.

## 2. Methods

### 2.1 Study design and data source

This study was part of a deep-insight visible neural network (DI-VNN) project. It applied an algorithm to predict several medical conditions, compared to other statistical and computational machine learning algorithms. Ethical review was exempted by the Taipei Medical University Joint Institutional Review Board (TMU-JIRB no.: N202106025).

There were two types of prediction models subsequentially developed in this study (Figure 1): (1) surrogate transcriptome models that derived each gene expression of a tissue type in the maternal-fetal interface from expressions of genes in maternal blood and (2) prediction models for any-onset preeclampsia using the surrogate transcriptome compared to that using the maternal blood transcriptome. The first type was to predict a condition that is occurring, i.e., a diagnostic prediction task; thus, we used a cross-sectional design. Meanwhile, the second type was to predict a condition in advance, i.e., a prognostic prediction task; thus, we used a prospective cohort design.

**Figure 1.**
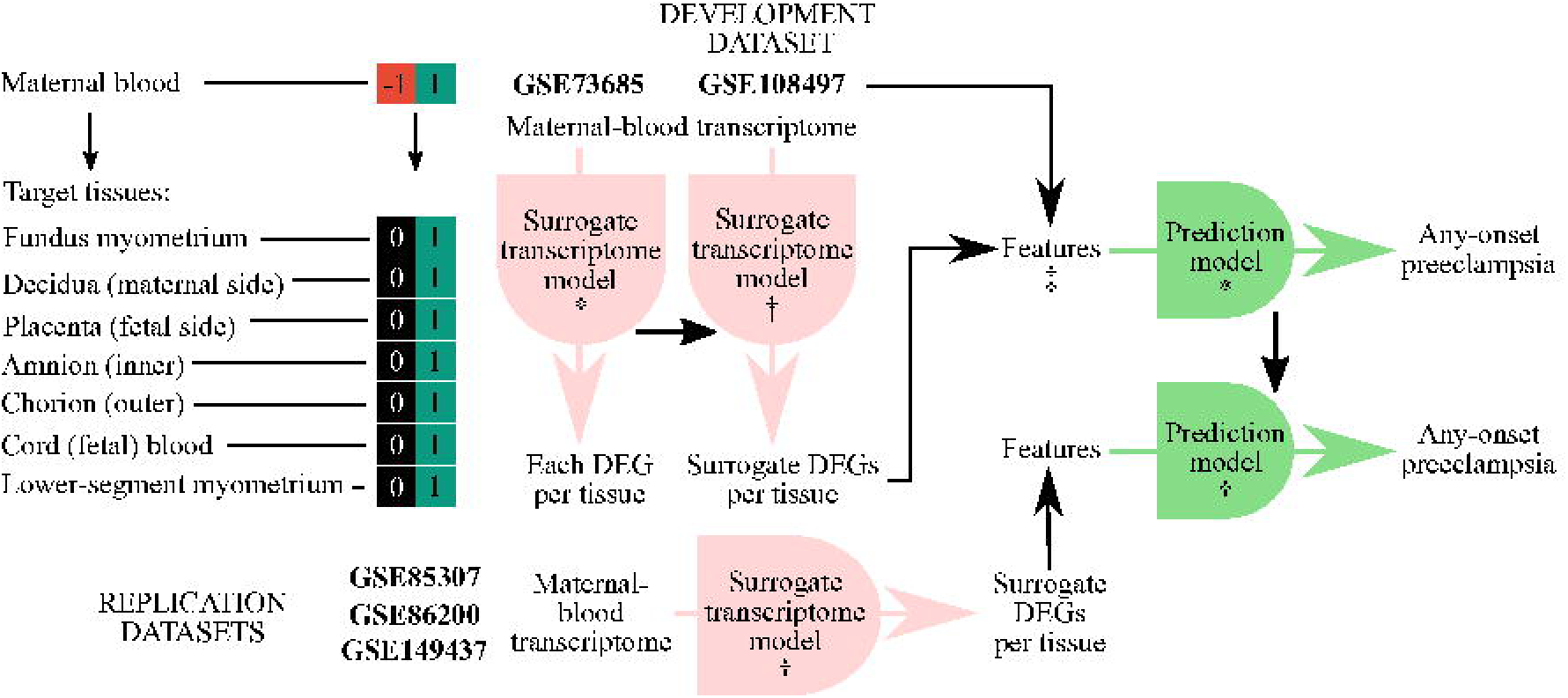
Predictive modeling pipeline. *, developed model; †, applied model; ‡, two models were developed using either the maternal-blood transcriptome or blood-derived surrogate; DEG, differentially expressed gene.

We utilized microarray datasets in the gene expression omnibus (GEO), a public functional genomics data repository (Table 1; see Data Availability) [22, 23]. For the derivation dataset of the surrogate transcriptome, we utilized gene expressions in multiple tissue samples from a healthy subject taken at the same time (*n*=183 samples; *n*=136 pairwise samples; GSE73685; total RNA extraction; GPL6244 Affymetrix Human Gene 1.0 ST Array) [24]. For the development dataset of the predictive modelling, we utilized gene expressions in maternal blood, including both EOPE and LOPE (*n*=512; GSE108497; total RNA extraction; GPL10558, Illumina Human HT-12 V4.0 expression beadchip) [25]. The prediction models were evaluated using the development dataset and those for replication: (1) an experimental dataset of a randomized controlled trial of vitamin D (25-hydroxyvitamin D [25OHD]) supplementation at up to 23 weeks’ gestation to prevent either EOPE or LOPE (*n*=157; GSE85307; total RNA extraction; GPL6244, Affymetrix Human Gene 1.0 ST Array) [26]; (2) an experimental dataset similar to the first one but using a different microarray platform with additional matched samples at 32 to 40 weeks’ gestation, and unspecified preeclampsia (*n*=60; GSE86200; total RNA extraction; GPL10558, Illumina Human HT-12 V4.0 expression beadchip) [27]; and (3) an observational dataset of a prospective cohort of pregnant women with EOPE and other conditions with shared pathophysiological derangement, including one unobserved in the development dataset (*n*=442; GSE149437; total RNA extraction; GPL28460, Affymetrix Human Transcriptome Array 2.0) [28].

**Table 1.** Derivation, development, replication, and coronavirus disease 2019 (COVID-19) datasets

| Outcome | Gestational age (weeks) |  |  |  | Total |
| --- | --- | --- | --- | --- | --- |
|  | <16 | 16~23 | 24~31 | 32~40 |  |
| Surrogate transcriptome model |  |  |  |  |  |
| GSE73685 (pairwise samples) – derivation dataset |  |  |  |  | 136 |
| Fundus myometrium vs. maternal blood |  |  |  |  | 19 |
| Decidua (maternal side) vs. maternal blood |  |  |  |  | 21 |
| Placenta (fetal side) vs. maternal blood |  |  |  |  | 14 |
| Amnion (inner) vs. maternal blood |  |  |  |  | 20 |
| Chorion (outer) vs. maternal blood |  |  |  |  | 20 |
| Cord (fetal) blood vs. maternal blood |  |  |  |  | 18 |
| Lower-segment myometrium vs. maternal blood |  |  |  |  | 22 |
| Excluded (technical outliers) |  |  |  |  | 2 |
| Prediction model |  |  |  |  |  |
| GSE108497 – development dataset (no intervention) |  |  |  |  | 512 |
| Normal (nonevent) | 75 | 69 | 73 | 68 | 285 |
| Isolated fetal growth restriction (small gestational age) (nonevent) | 6 | 7 | 5 | 3 | 21 |
| Early-onset preeclampsia (event) | 8 | 8 | 6 | 0 | 22 |
| Late-onset preeclampsia (event) | 4 | 3 | 3 | 3 | 13 |
| Excluded (technical outliers; outcome with extremely underrepresented gestational age, i.e. $n=1$) | | | | | 171 |
| GSE85307 – replication dataset (vitamin D +/-) |  |  |  |  | 157 |
| Normal (nonevent) | 64 | 44 | 0 | 0 | 108 |
| Early-onset preeclampsia (event) | 28 | 13 | 0 | 0 | 41 |
| Late-onset preeclampsia (event) | 4 | 2 | 0 | 0 | 6 |
| Excluded (technical outliers) |  |  |  |  | 2 |
| GSE86200 – replication dataset (vitamin D +/-) |  |  |  |  | 60 |
| Normal (nonevent) | 17 | 7 | 0 | 24 | 48 |
| Preeclampsia (event) | 5 | 1 | 0 | 6 | 12 |
| Excluded (technical outliers) |  |  |  |  | 1 |
| GSE149437 – replication dataset (no intervention) |  |  |  |  | 442 |
| Normal (nonevent) | 0 | 0 | 0 | 20 | 20 |
| Spontaneous preterm delivery (nonevent) | 25 | 45 | 62 | 30 | 162 |
| Preterm premature rupture of membranes (nonevent) | 26 | 52 | 73 | 36 | 187 |
| Early-onset preeclampsia (event) | 11 | 23 | 23 | 9 | 66 |
| Excluded (technical outliers) |  |  |  |  | 7 |
| GSE177477 – COVID-19 dataset |  |  |  |  | 47 |
| Uninfected controls (nonevent) |  |  |  |  | 18 |
| Asymptomatic cases (event) |  |  |  |  | 18 |
| Mild cases (event) |  |  |  |  | 3 |
| Severe cases (event) |  |  |  |  | 8 |
| Excluded (technical outliers) |  |  |  |  | 0 |

The derivation, development, and replication datasets were all collected before the worldwide COVID-19 pandemic (Figure A.1). For the COVID-19 dataset, we utilized another microarray dataset to predict cases infected by COVID-19 (*n*=47; GSE177477; total RNA extraction; GPL23159, Affymetrix Clariom S Assay with Pico Assay), consisting of: (1) uninfected controls (*n*=18); (2) asymptomatic cases (*n*=18); (3) mild, symptomatic cases (*n*=3); and (4) severe, symptomatic cases (*n*=8) [29]. We also utilized a validated gene set to predict any-onset preeclampsia during the pandemic (*n*=404; GSE192902; total RNA extraction; GPL24676, Illumina NovaSeq 6000) [30]. The gene set was well-replicated, especially for predicting preeclampsia, as validated by an independent dataset from another study, which was collected from February 2017 to January 2019 and from April 2017 to July 2018. Predicting preeclampsia using any transcripts in the gene set might be shared with that of COVID-19 infection. But, this dataset also allowed us to distinguish if the shared prediction (if any) was because (1) a possibility that the discovery dataset included pregnant women with undiagnosed, asymptomatic COVID-19 or (2) a methodological limitation of identifying a unique blood biomarker under endothelial dysfunction. Although shared sFlt-1 predictions were those between preeclampsia and severe COVID-19 [4, 5], we chose all of the conditions under COVID-19 infection, including asymptomatic cases. It might likely be a false positive for preeclampsia if the prognostication is not in conjunction with a COVID-19 test result. A doctor would unlikely order a COVID-19 test if none of the indications was identified, except those tests that are universally applied to all the pregnant women in a healthcare facility.

### 2.2 Derivation of the maternal-fetal interface transcriptome from maternal blood

A standard preprocessing pipeline of microarray data was applied (see Appendix A). This included background correction, probe set normalization, removal of technical outliers (Table 1), removal of low-expressed probe sets, gene annotation, summarization from probe sets to genes, and selection of common genes among all the microarray platforms. A differential expression analysis was conducted with batch-effect removal using a singular value approximation. In the analysis, a moderated *t*-statistic was applied using pairwise samples of maternal blood and each tissue at the maternal-fetal interface. This was subsequentially followed by the Benjamini-Hochberg multiple-testing correction with a maximum false discovery rate (FDR) of 0.05 to determine if a gene was differentially expressed. Therefore, we identified differentially expressed genes (DEGs) for each tissue type at the maternal-fetal interface compared to those in maternal blood, and computed average expressions in maternal blood and/or each of the tissue types.

We developed a surrogate transcriptome model for predicting each individual-level DEG of a tissue type at the maternal-fetal interface, but only considered genes that were differentially expressed in that tissue type, using gene expression in maternal blood as candidate predictors. We defined the predicted outcome to reflect individual-level DEGs that considerably differed from the gene expression distribution in the tissue type against that in maternal blood, but not always extremely different (e.g. >95^th^ percentile). Candidate predictors were also standardized using average expression numbers derived from the differential analyses. A definition of the outcome and standardization of the candidate features are described (see Appendix A).

A surrogate transcriptome model was only developed for a gene with greater than or equal to three instances for the minority outcome and a minimum of two candidate predictors. Considering the tradeoff between the number of genes fulfilling the aforementioned criteria and the risk of bias due to a small sample size, we applied a protocol to reduce the number of candidate predictors without leaking the outcome information to prevent overfitting, as described previously [31]. This resulted in cross-validated principal components (PCs) which were used as candidate predictors. We applied a logistic regression with regularization, which was an elastic net regression, and subsequentially recalibrated with either a linear regression model or a general additive model using locally weighted scatterplot smoothing (GAM-LOESS). Each model estimated a probability of 0 to 1 of how likely a gene of a tissue type in an individual was differentially expressed compared to that of maternal blood.

### 2.3 Development and replication of a prediction model for any-onset preeclampsia

We argue that maternal blood DEGs cannot be generalized predictors for preeclampsia since these may be misleading due to abrupt changes in endothelial dysfunction which varies widely among pregnant women with different comorbidities. But, if we take the maternal blood transcriptome into account, regardless of whether a gene’s expression is differential to preeclampsia, we may obtain a biological signal unique to this condition, if it represents gene expression in tissues at the maternal-fetal interface, at least to some extent. To support this argument, we compared predictive performances using the development and replication datasets between (1) models that used the maternal-blood transcriptome and (2) models that used the blood-derived surrogate transcriptome of maternal-fetal interface tissues.

For the first type of model, we used the transcriptome of maternal blood as candidate predictors. We conducted a differential expression analysis to independently identify DEGs between preeclampsia and non-preeclampsia in the development and replication datasets. The analysis pipeline was the same as that for deriving the surrogate transcriptome, but the comparison was not pairwise. We only used DEGs as the maternal-blood transcriptome for the first type of model. The transcriptome should also intersect with that used as candidate predictors for deriving the surrogate transcriptome. If this type of model could not replicate the predictive performance, then this implies that maternal blood DEGs of preeclampsia likely reflect endothelial dysfunction which varies widely among pregnant women with different comorbidities. In addition, we examined overlapping DEGs among these datasets to identify common genes for an exploratory analysis.

For the second type of model, we used the blood-derived surrogate transcriptome of the maternal-fetal interface as candidate predictors. The surrogate transcriptome of each tissue in the maternal-fetal interface was derived from the maternal-blood transcriptome using surrogate models, as described in the previous section. But, instead of the maternal-blood transcriptome in derivation dataset, we used those in datasets for developing and replicating a prediction model for any-onset preeclampsia. Before deriving the surrogate transcriptome, quantile-to-quantile normalization followed by standardization of candidate predictors was applied for each of those datasets based on average expressions of genes in each of the tissues of the derivation dataset. Since the surrogates have different accuracies among genes to predict the true transcriptome, we applied different weights among genes of the surrogate transcriptome by multiplying the expression probability by Matthew’s correlation coefficient (MCC). Its value ranges between -1 and 1, in which 1 means perfect accuracy, 0 means poor accuracy, and -1 means inverted accuracy. We normalized the multiplication results into values from 0 to 1.

In addition to candidate predictors, modeling algorithms may also contribute to the predictive performance. We applied several machine learning algorithms to develop a prediction model using each set of candidate predictors: (1) principal-component (PC)-elastic net regression (ENR); (2) PC-random forest (RF); (3) PC-gradient boosting machine (GBM); and (4) deep-insight visible neural network (DI-VNN). These models were also recalibrated by either a linear regression model or GAM-LOESS. The analysis pipeline for comparison among these algorithms was described previously [32]. But, we excluded irrelevant procedures and conducted external validation to replicate the models using independent datasets instead of excluding samples by either simple or stratified random sampling. Before replicating the models that used the transcriptome of maternal blood, we applied quantile-to-quantile normalization to gene expressions of the three replication datasets based on average expressions of genes in the development dataset.

### 2.4 Emulation of potential RT-qPCR-based blood biomarkers for any-onset preeclampsia

We also compared different methods of predictor discovery for emulating potential blood biomarkers to propose low-cost predictions of any-onset preeclampsia in clinical practice, as those using a reverse-transcription quantitative polymerase chain reaction (RT-qPCR). Blood biomarkers were from: (1) the best model among those using either the maternal-blood transcriptome or blood-derived surrogate transcriptome of maternal-fetal interface tissues; (2) DEGs of the development dataset with either very low or high expression (absolute log_2_ [fold change] of >2); (3) DEGS of the development dataset but not in both the development and replication datasets without an intervention; (4) DEGs of both the development and replication datasets without an intervention; and (5) a validated gene set from a previous study [30] for predicting any-onset preeclampsia, including a period during the COVID-19 pandemic. From the latter, we could only use 10 of 18 genes in the validated gene set, because these genes were available in all the derivation, development, replication, and COVID-19 datasets. The genes were *CAMK2G*, *DERA*, *KIAA1109*, *LRRC58*, *NDUFV3*, *NMRK1*, *PYGO2*, *RNF149*, *TFIP11*, and *TRIM21*.

We used combinations with one to five members from the list of biomarkers for each method. The number of members was chosen to achieve low-cost predictions. However, the number of members in each combination might not be maximized, since the number of combinations expands exponentially depending on the number of biomarkers in the list.

To emulate gene expression values by the RT-qPCR, gene expressions were standardized (i.e., using Z-scores) with the average and standard deviation (SD) calculated from the development dataset without outliers. These were defined as values of less than or more than 1.5 times the interquartile range, respectively, from the first or third quantile. Yet, none of the outliers were excluded. The emulation was conducted using a decision tree algorithm with the maximum depth depending on the number of biomarkers in only the development dataset.

### 2.5 Utilizing preeclampsia blood biomarkers for predicting COVID-19 infection

To ensure that biomarkers were unique to any-onset preeclampsia but not COVID-19 infection, we utilized the emulated blood biomarkers to predict COVID-19 infection as the event. The blood biomarkers were expected to acquire lower performance for predicting COVID-19 infection than that for predicting preeclampsia. Although a previous study demonstrated that sFlt-1, a well-known blood biomarker for preeclampsia, could predict severe cases of COVID-19 [4, 5], we could not reevaluate the finding in this study, because this biomarker is in a protein form. Instead, we included transcripts from the validated gene set as the fifth method of predictor discovery [30]. Since this gene set was discovered by a differential expression analysis during the COVID-19 pandemic, it is possible that the method falsely discovers blood biomarkers misled by endothelial dysfunction which is a key pathogenic mechanism shared between preeclampsia and COVID-19 infection [16, 17].

### 2.6 Statistical analysis

Bootstrapping 30 times was applied to infer the 95% confidence interval (CI) of the predictive performances of the prediction models and emulated blood biomarkers. The performance of a prediction model was measured by the area under receiver operating characteristics curve (AUROC) which reflects true positive and negative rates. The models should be well-replicated, which was an interval estimate of an AUROC of ≥0.5 and more than the average per combination of a dataset and a set of candidate predictors, in the development and replication datasets, particularly those without an intervention (i.e. GSE108497 and GSE149437). The best model was evaluated for each set of candidate predictors based on the AUROC of the development dataset.

For each method of predictor discovery, we computed the number of biomarker combinations that could predict any-onset preeclampsia but not COVID-19 infection. Specifically, the combination should fulfill these criteria: (1) the point estimate of the AUROC for predicting preeclampsia in the replication dataset without an intervention is between the interval estimate of that in the development dataset; (2) the point estimate of the AUROC for predicting COVID-19 infection is smaller or equal to the lower bound of the AUROC interval estimate for predicting preeclampsia in the development dataset; and (3) the lower bound of the AUROC interval estimate for predicting COVID-19 infection was not greater or equal to 0.5. We conducted a permutation test (500 iterations) for each method of predictor discovery. If the *P*-value was >0.05, then the biomarkers fulfilled the criteria by chance, i.e., the null hypothesis was accepted. Rejecting the null hypothesis meant that a method significantly discovered predictors that could predict any-onset preeclampsia but not COVID-19 infection. The best emulated biomarkers were taken from the significant method with the greatest number of biomarkers fulfilling the criteria.

We also conducted an exploration and reanalysis of low- and high-level information from databases of the GeneCards human genes (version 5.7; December 6, 2021),[33] the DIANA miRNA tissue expression (15,183 datasets; miRBase version 22) [34], and the STRING functional protein association network (version 11.5; latest update August 12, 2021) [35]. These were related to the best model, especially the best emulated blood biomarkers. The analysis codes and details, including versions, are being shared publicly to allow replication of this study (see Code Availability). All analyses were conducted using R except for retrieving the annotation. Webpages of the retrieved information from the GeneCards and STRING, which were reserved at the time of accession in the Internet Archive and can be re-accessed via its Wayback Machine (see Appendix A). For DIANA and STRING, we downloaded the datasets of the information that was retrieved for this study to be shared in Appendices A and B and the analysis codes and details.

## 3. Results

### 3.1 Subject characteristics

Only pregnancy outcome data were publicly shared at the individual level by the original study which collected the derivation dataset (Table 2). While the tissues were obtained during a cesarean delivery, all deliveries with or without labor were represented, with either preterm or term delivery. These also included preterm deliveries with prelabor rupture of the membrane (PROM). The derivation dataset did not publicly share which individuals were preeclamptic (*n*=3) among the pregnant women with a preterm delivery but without labor (*n*=10), as reported in the publication [24]. The COVID-19 dataset did not publicly share subject characteristics for either uninfected or asymptomatic COVID-19 individuals, except for sex.

**Table 2.** Subject characteristics of derivation, development, and replication datasets.

| Variable | Nonevent | Event | P-value |
| --- | --- | --- | --- |
| **Derivation dataset** |  |  |  |
| **GSE73685 (*n*, %) \*** | **134 (100)** |  |  |
| Preterm with labor (*n*, %) | 9 (6.72) |  |  |
| Preterm without labor (*n*, %) | 30 (22.39) | † |  |
| Preterm PROM with labor (*n*, %) | 11 (8.21) |  |  |
| Preterm PROM without labor (*n*, %) | 11 (8.21) | † |  |
| Term with labor (*n*, %) | 27 (20.15) |  |  |
| Term without labor (*n*, %) | 46 (34.33) |  |  |
| **Development dataset** |  |  |  |
| **GSE108497 (*n*, %)** | **306 (100)** | **35 (100)** |  |
| Maternal age at collection (year, SD) | 31 (5) | 31 (4) | >.05 |
| Gestational age at collection (week, SD) | 23 (10) | 20 (8) | >.05 |
| Ethnicity of Hispanic or Latino: |  |  |  |
| No (*n*, %) | 261 (85.29) | 25 (71.4) | (reference) |
| Yes (*n*, %) | 45 (14.71) | 10 (28.6) | .039 |
| Systemic lupus erythematosus: |  |  |  |
| No (*n*, %) | 147 (48.04) | 0 (0) | (reference) |
| Yes (*n*, %) | 159 (51.96) | 35 (100) | >.05 |
| **Replication datasets** |  |  |  |
| **GSE85307 (*n*, %)** | **108 (100)** | **47 (100)** |  |
| Maternal age at collection (year, SD) | 27 (5) | 26 (5) | >.05 |
| Gestational age at collection (week, SD) | 14 (3) | 14 (3) | >.05 |
| Ethnicity: |  |  |  |
| White (*n*, %) | 42 (38.89) | 17 (36.2) | (reference) |
| Black or African American (*n*, %) | 59 (54.63) | 22 (46.8) | >.05 |
| Asian (*n*, %) | 2 (1.85) | 2 (4.3) | >.05 |
| American Indian or Alaska (*n*, %) | 0 (0.00) | 3 (6.4) | >.05 |
| Other (*n*, %) | 5 (4.63) | 3 (6.4) | >.05 |
| Body-mass index (kg/m<sup>2</sup>, SD) | 27.68 (7.33) | 31.20 (8.00) | .010 |
| Asthma: |  |  |  |
| No (*n*, %) | 65 (60.19) | 27 (57.5) | (reference) |
| Yes (*n*, %) | 43 (39.81) | 20 (42.6) | >.05 |
| Vitamin D baseline (ng/mL whole blood, SD) | 27.68 (7.33) | 31.20 (8.00) | .010 |
| **GSE86200 (*n*, %) \*** | **48 (100)** | **12 (100)** |  |
| Maternal age at enrollment (year, SD) | 25 (6) | 24 (5) | >.05 |
| Gestational age at enrollment (week, SD) | 14 (3) | 13 (3) | >.05 |
| Ethnicity: |  |  |  |
| Caucasian, Non-Hispanic (*n*, %) | 12 (25) | 0 (0) | (reference) |
| Black or African American (*n*, %) | 36 (75) | 12 (100) | >.05 |
| Fetal sex: |  |  |  |
| Female (*n*, %) | 28 (58) | 2 (17) | (reference) |
| Male (*n*, %) | 20 (42) | 10 (83) | >.05 |
| Vitamin D at enrollment (nmol/L whole blood, SD) ‡ | 51.4 (26.6) ¶ | 30.8 (9.6) ¶ | >.05 |
| Vitamin D at third trimester (nmol/L whole blood, SD) ‡ | 84.9 (34.0) ¶ | 63.5 (47.1) ¶ | >.05 § |
| **GSE149437 (*n*, %) \|\|** | **369 (100)** | **66 (100)** |  |
| Gestational age at collection (week, SD) | 25 (8) | 22 (7) | .008 |

\*, number of pairwise samples, of which those in GSE86200 are shown as unpaired numbers (i.e., doubling); †, preeclampsia in three of 10 pregnant women with preterm without labor [24], but the information of which samples were undisclosed; ‡, 1 nmol/L = 0.2885 ng/mL; ¶, nonevent (*n*=24) and event (*n*=6); §, significantly differ in the parent study of Al-Garawi, et al (2016), which had a larger sample size (*n*=806) [84]; ||, number of samples from both the same and different subjects; PROM, prelabor rupture of the membranes; SD, standard deviation.

Among the development and replication datasets (Table 2), only gestational age in the replication dataset without an intervention differed between the event and nonevent groups. This dataset also did not report maternal age or ethnicity. Maternal ages in the replication datasets with an intervention were younger compared to those in the development dataset. Ethnicity only differed in the development dataset for Hispanic or Latino women. Other replication datasets with an intervention only reported non-Hispanic/Latino ethnicities. Vitamin D intervention data were not shared publicly in the datasets, but vitamin D blood levels were reported at the baseline or enrollment. Only one of two replication datasets reported vitamin D blood levels in the third trimester. Those did not differ between events and nonevents in this dataset, which was one for microarray analysis; however, the dataset was only a subset of a larger dataset in the parent study. Vitamin D blood levels in the third trimester significantly differed in the parent study [27]; thus, this replication dataset likely has smaller power to detect differences in vitamin D blood levels in the third trimester which was the time after a vitamin D intervention. For the COVID-19 dataset, we identified eight of 23 females and 21 of 30 males in the publication (*n*=53)^1^ who were diagnosed with COVID-19 infection in the dataset. Of 24 uninfected individuals,^2^ that were reported in the publication [29], this dataset did not share six of them, leaving only 47 subjects.

### 3.2 Blood-derived surrogate transcriptome of the maternal-fetal interface

MCCs did not significantly differ by interval estimates for predicting individual-level DEGs among those in maternal-fetal interface tissues (Figure 2). The surrogate models had neither poor (MCC=0) nor inverted (MCC<0) accuracy. Of the DEGs that could be predicted at the individual level (Figure 2; Table 3), the placenta had the smallest number (*n*=442), while the decidua had the largest number (*n*=967). Meanwhile, the smallest and largest numbers of DEGs (Table 3) were respectively found in the decidua (*n*=6704) and lower-segment myometrium (*n*=7574).

**Figure 2.**
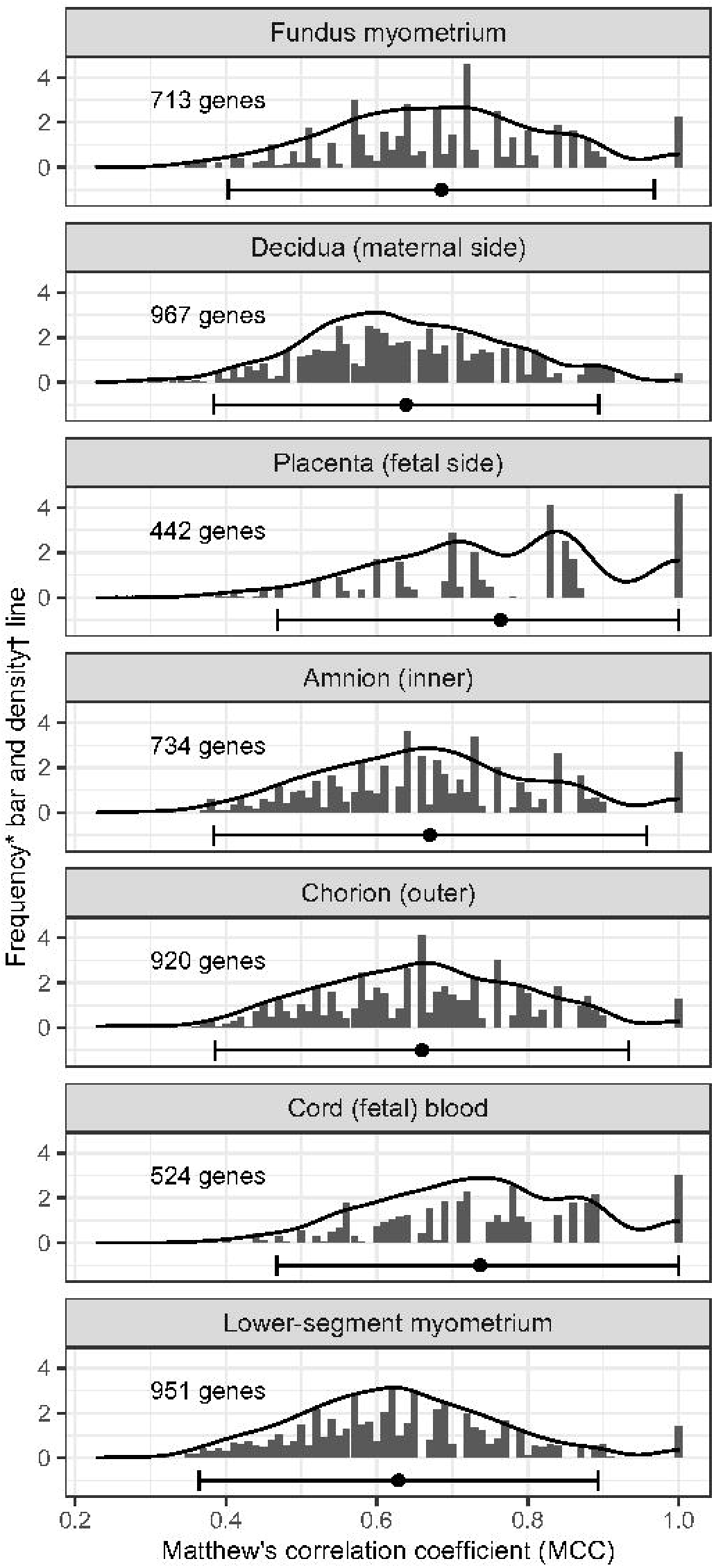
Distribution of weights used to adjust the gene expression probability. The weight was determined by Matthew’s correlation coefficient (MCC) and rounded to two decimal places for binning MCCs. *, ratio of the number of genes per MCC bin and the average number per tissue; †, probability of distribution.

**Table 3.** Surrogate transcriptome among differentially expressed genes (DEGs).

| Target tissue (GSE73685) | Proportion of surrogate transcriptome |  |  |  | Non-DEG | Total |
| --- | --- | --- | --- | --- | --- | --- |
|  | Log<sub>2</sub> FC of DEG (target tissue vs. maternal blood) |  |  |  |  |  |
|  | >2 | 0 to 2 | <0 to -2 | <-2 |  |  |
| Fundus myometrium (*n/N*, %) | 2/489<br>(0.41) | 339/3383<br>(10.02) | 370/3141<br>(11.78) | 2/512<br>(0.39) | 0/1695<br>(0) | 713/9220<br>(7.73) |
| Decidua (maternal side) (*n/N*, %) | 8/239<br>(3.35) | 466/3133<br>(14.87) | 491/3159<br>(15.54) | 2/173<br>(1.16) | 0/2516<br>(0) | 967/9220<br>(10.49) |
| Placenta (fetal side) (*n/N*, %) | 0/393<br>(0) | 193/2910<br>(6.63) | 249/2902<br>(8.58) | 0/573<br>(0) | 0/2442<br>(0) | 442/9220<br>(4.79) |
| Amnion (inner) (*n/N*, %) | 3/413<br>(0.73) | 331/3521<br>(9.4) | 385/2997<br>(12.85) | 15/532<br>(2.82) | 0/1757<br>(0) | 734/9220<br>(7.96) |
| Chorion (outer) (*n/N*, %) | 14/386<br>(3.63) | 448/3185<br>(14.07) | 451/2835<br>(15.91) | 7/465<br>(1.51) | 0/2349<br>(0) | 920/9220<br>(9.98) |
| Cord (fetal) blood (*n/N*, %) | 1/36<br>(2.78) | 285/1902<br>(14.98) | 238/1886<br>(12.62) | 0/4<br>(0) | 0/5392<br>(0) | 524/9220<br>(5.68) |
| Lower-segment myometrium (*n/N*, %) | 7/444<br>(1.58) | 453/3367<br>(13.45) | 482/3359<br>(14.35) | 9/404<br>(2.23) | 0/1646<br>(0) | 951/9220<br>(10.31) |

FC, fold change; *n*, number of genes predicted by the surrogate transcriptome model; *N*, number of genes in the differential expression analysis.

Proportions of the tissue transcriptome that could be predicted from that of maternal blood (Table 3) were from 4.79% (placenta) to 10.49% (decidua). The placenta had the smallest proportion of the surrogate transcriptome, and none of the DEGs had an absolute value of >2 log_2_ [fold change]. This implied that only a small proportion of the transcriptome of maternal-fetal interface tissues could be represented by that of maternal blood, especially the placenta transcriptome.

### 3.3 A prediction model for any-onset preeclampsia using the surrogate transcriptome

To develop comparator prediction models, we only used DEGs (*n*=924) based on the development dataset (Table 4), from the blood transcriptome which intersected with those used as candidate predictors for deriving the surrogate transcriptome (*n*=7524). After developing the prediction models using the blood transcriptome with several algorithms (Figure 3), none of the predictive performances were well-replicated, although the average was higher than that using the blood-derived surrogate transcriptome based on the development dataset. In addition, overlapping DEGs (Table 4) were only found between the development and replication datasets without an intervention (*n*=25).

**Figure 3.**
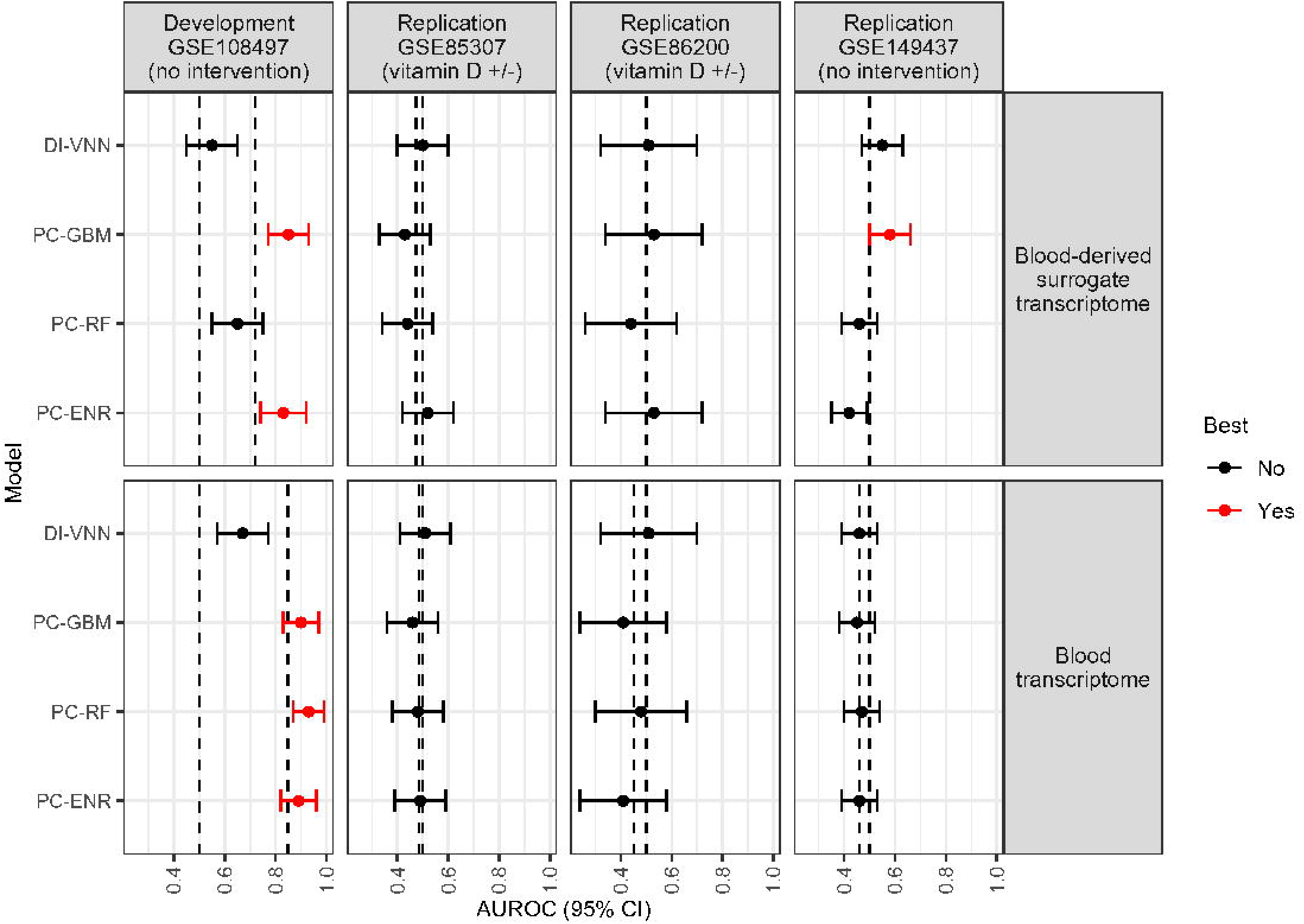
Predictive performance between models using the maternal-blood transcriptome and blood-derived surrogate in all datasets. Dashed lines show the area under receiver operating characteristics curve (AUROC) of 0.5 and the average per dataset among models using the same set of candidate predictors. The best model was evaluated in each set of candidate predictors by the AUROC. If the AUROC interval was ≥0.5 and more than the average in the development and replication datasets, particularly those without an intervention (i.e., vitamin D supplementation), the model was well-replicated. CI, confidence interval; DI-VNN, deep-insight visible neural network; ENR, elastic net regression; GBM, gradient boosting machine; PC, principal component; RF, random forest.

**Table 4.**
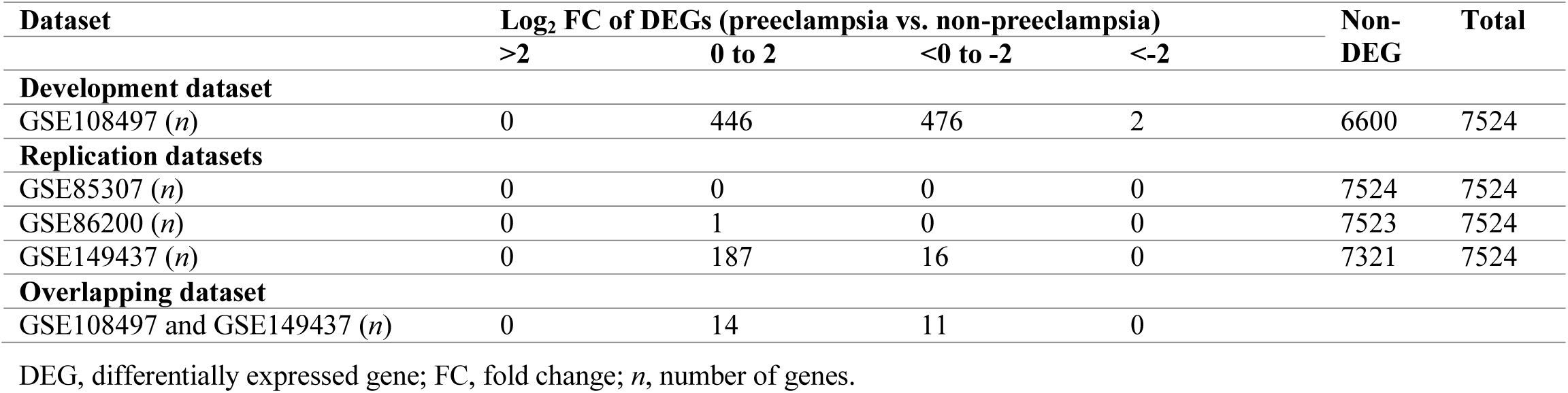
Differential expression independently among the datasets.

| Dataset | Log<sub>2</sub> FC of DEGs (preeclampsia vs. non-preeclampsia) |  |  |  | Non-DEG | Total |
| --- | --- | --- | --- | --- | --- | --- |
|  | >2 | 0 to 2 | <0 to -2 | <-2 |  |  |
| **Development dataset** |  |  |  |  |  |  |
| GSE108497 (*n*) | 0 | 446 | 476 | 2 | 6600 | 7524 |
| **Replication datasets** |  |  |  |  |  |  |
| GSE85307 (*n*) | 0 | 0 | 0 | 0 | 7524 | 7524 |
| GSE86200 (*n*) | 0 | 1 | 0 | 0 | 7523 | 7524 |
| GSE149437 (*n*) | 0 | 187 | 16 | 0 | 7321 | 7524 |
| **Overlapping dataset** |  |  |  |  |  |  |
| GSE108497 and GSE149437 (*n*) | 0 | 14 | 11 | 0 |  |  |

DEG, differentially expressed gene; FC, fold change; *n*, number of genes.

Meanwhile, we found a well-replicated predictive performance of one of the prediction models using the blood-derived surrogate transcriptome (Figure 3) in the development and replication datasets without an intervention. This applied the PC-GBM which was also applied for one of the prediction models using the blood transcriptome. There were 108 predictors from the surrogate transcriptome in any of the tissues, which were derived from 5897 predictors from the blood transcriptome.

### 3.4 Potential blood biomarkers unique to any-onset preeclampsia but not COVID-19 infection

Nevertheless, a prediction model that uses 5897 predictors is costly in clinical settings; thus, we needed to choose a few predictors of the maternal blood transcriptome to predict any-onset preeclampsia. After our data analysis showed that the best model was the PC-GBM, we determined how to plausibly choose a few of the predictors from this model. But, an exploratory approach should not be used to avoid confirmatory bias by investigators; thus, we did exhaustive comparisons of a few predictors using decision trees, as applied for other methods of predictor discovery (see Subsection 2.4).

Since we needed to choose predictors that represented the transcriptome of all maternal-fetal interface tissues, blood-derived predictors were chosen if these were included in predictors with the top one to 20 absolute values of average weights, that predicted the surrogate transcriptome of each tissue type. Predictors with the top one to five values in all the tissue types were subsequentially chosen; thus, we developed 20 × 5 decision trees. None of the selected predictors were genes in the DEGs of the development dataset, which meant that the most important predictors that predicted the surrogate transcriptome in each tissue and all tissues were not extremely expressed genes in maternal blood.

Eventually, we chose the best method of predictor discovery based on the significantly greatest number of eligible biomarkers (Table 5), which was intended to find those for predicting any-onset preeclampsia but not COVID-19 infection (see Subsection 2.6). Only the blood-derived surrogate transcriptome by the PC-GBM significantly discovered eligible biomarkers (*n*=3/100, 3.0%; *P*=.036). These were combined from different candidate predictors corresponding to the surrogate transcriptome in different tissues at the maternal fetal interface.

**Table 5.** Number of biomarkers for any-onset preeclampsia but not severe coronavirus disease 2019 (COVID-19).

| Method of predictor discovery | Eligible biomarkers | *P*-value |
| --- | --- | --- |
| **Blood-derived surrogate transcriptome** |  |  |
| PC-GBM, top 1 to 20 of surrogate genes, top 1 to 5 of the blood genes (*n/N*, %) | 3/100 (3.0%) | .036 |
| **Blood transcriptome** |  |  |
| DEGs of GSE108497 *, absolute log<sub>2</sub> fold change >2 (*n/N*, %) | 0/1 (0%) | .018 |
| DEGs of GSE108497 * and GSE149437 †, 1 to 2 combinations from 25 DEGs (*n/N*, %) | 3/325 (0.09%) | >.05 |
| DEGs of GSE108497 but not in both GSE108497 * and GSE149437 †, each from 899 DEGs (*n/N*, %) | 13/899 (1.45%) | >.05 |
| DEGs of a recent study (18 genes), 1 to 2 combinations from 10 DEGs in GSE108497 * and GSE177477 ‡ | 0/55 (0.0%) | >.05 |

\*, the development dataset; †, the replication dataset without an intervention; ‡, COVID-19 dataset; DEG, differentially expressed gene; PC-GBM, principal-component gradient boosting machine.

The three combinations resulted in final decision trees, each of which consisted of the same predictors, i.e., *ITGA5*, *P2RX7*, and *IRF6*, but the trees had different splitting cutoffs for each predictor. We chose the tree developed with the least number of candidate predictors, which was that using the criteria of the top three surrogate genes and the top two blood genes (Figure 4). Transcript of *ITGA5* with a Z-score of ≥1.1 (terminal branch A) defined the majority of the predicted events (73.70%) among preeclampsia samples in the development dataset, but only a minority of predicted events (6.34%) among positives in the COVID-19 dataset. Otherwise, to define predicted events in the development dataset, we only needed a subsequent measurement of the *IRF6* transcript with a Z-score of ≥-0.73 (terminal branches B and C). This was regardless of the *P2RX7* transcript. For predicted events in the COVID-19 datasets, none was defined by the *P2RX7* transcript with a Z-score of <0.13 (terminal branch C), but this defined a minority of predicted events (9.87%) among preeclampsia samples in the development dataset. If we only used samples with either EOPE or LOPE in the development dataset, proportions of predicted events were respectively shifted away or toward terminal branch A (Figure 4).

**Figure 4.**
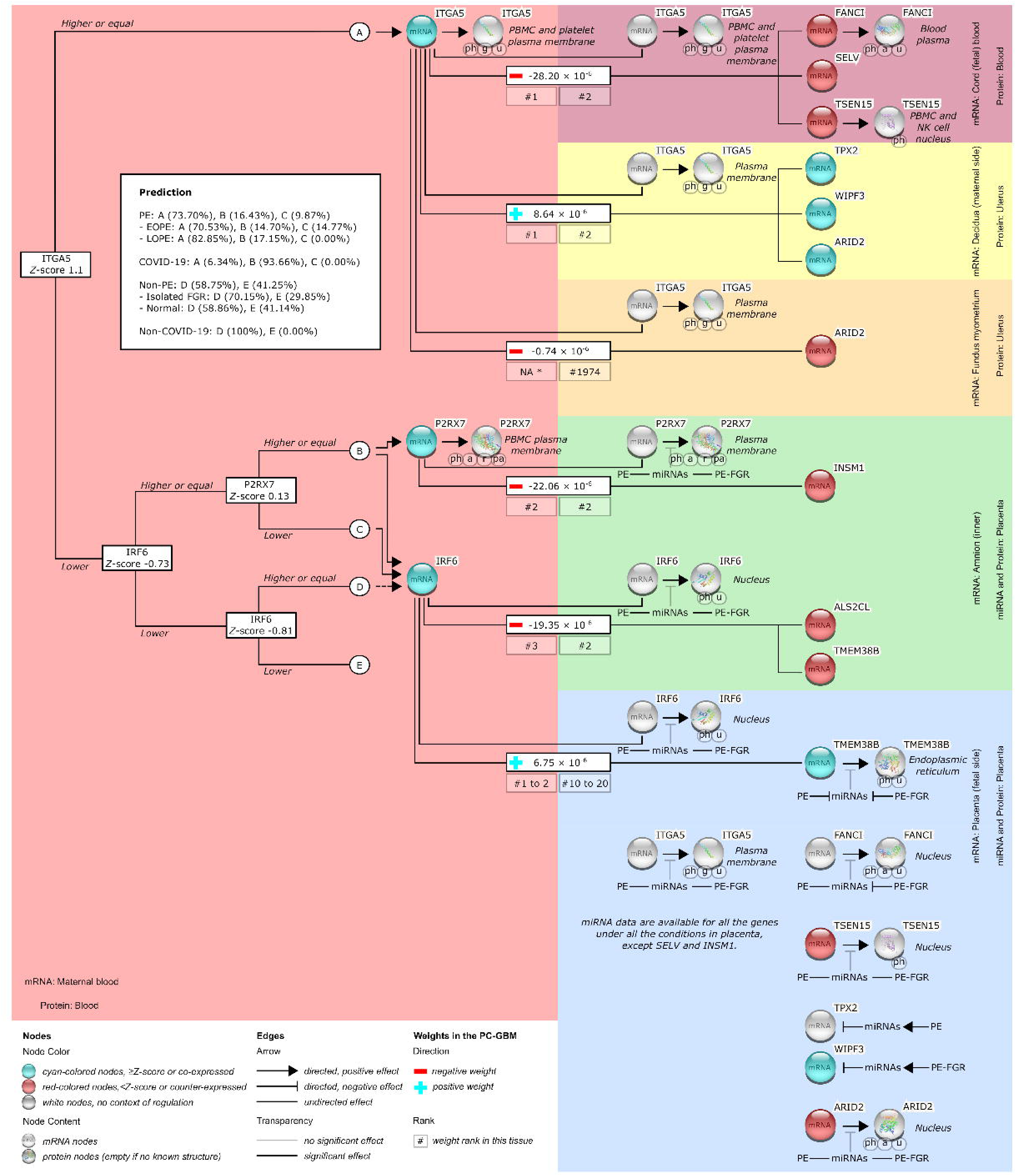
Emulation of the most predictive biomarkers from the principal component-gradient boosting machine (PC-GBM). The number is the standardized value of the splitting biomarker. A dashed**-**line arrow from node D to the IRF6 mRNA node is applied only if P2RX7 is not measured. *, not fulfilling the criteria (i.e., top one to 20 of surrogate genes and top one to 5 of blood genes); a, acetylation; EOPE, early-onset preeclampsia (PE); FGR, fetal growth restriction; g, glycosylation; LOPE, late-onset PE, pa, palmitoylation; PE, preeclampsia; ph, phosphorylation; r, ribosylation; u, ubiquitination.

None of the predicted events was defined by terminal branch C if we only used samples with LOPE in the development dataset. If we only used samples with either normal or isolated FGR in the development dataset, proportions of the predicted nonevents were respectively shifted away or toward terminal branch D (Figure 4). The predicted nonevents were less (29.85% vs. 41.25%) defined by terminal branch E if we only used samples with isolated FGR in the development dataset, compared to those with all non-preeclamptic conditions. Therefore, terminal branches A, B, C, D, and E (Figure 4) respectively tended to predict LOPE, COVID-19 infection or any PE, EOPE, more-isolated FGR than a normal condition, and more-normal condition than isolated FGR.

### 3.5 Post-analysis justification for the biological relevance of the best potential biomarkers

In the PC-GBM (Figure 4), the *ITGA5* transcript in maternal blood was used to predict surrogate transcripts of: (1) *FANCI*, *SELENOV* (*SELV*), and *TSEN15* in cord blood; (2) *TPX2*, *WIPF3*, and *ARID2* in decidua; and (3) *ARID2* in the fundus myometrium. But, the *ITGA5* transcript in maternal blood, that predicted the surrogate transcript of *ARID2* in fundus myometrium, did not fulfill the criteria to be included in the emulated biomarkers (ranked within top one to 20 surrogate genes and top one to five blood genes). The *P2RX7* transcript in maternal blood was used to predict the *INSM* surrogate transcript in the amnion. Eventually, the *IRF6* transcript in maternal blood was used to predict the surrogate transcripts of: (1) *ALS2CL* and *TMEM38B* in the amnion; and (2) *TMEM38B* in the placenta. But, the *IRF6* transcript in maternal blood, that predicted the surrogate transcript of TMEM38B in the placenta, was not ranked in top three of surrogate genes in this tissue.

To justify the biological relevance of these weights and cutoffs, we conducted an exploration and reanalysis of low- and high-level information from the databases, related to *ITGA5*, *P2RX7*, and *IRF6*, with the surrogate transcriptome. The biological relevance based on this information is elaborated (Figure 5; see Subsections 4.2).

**Figure 5.**
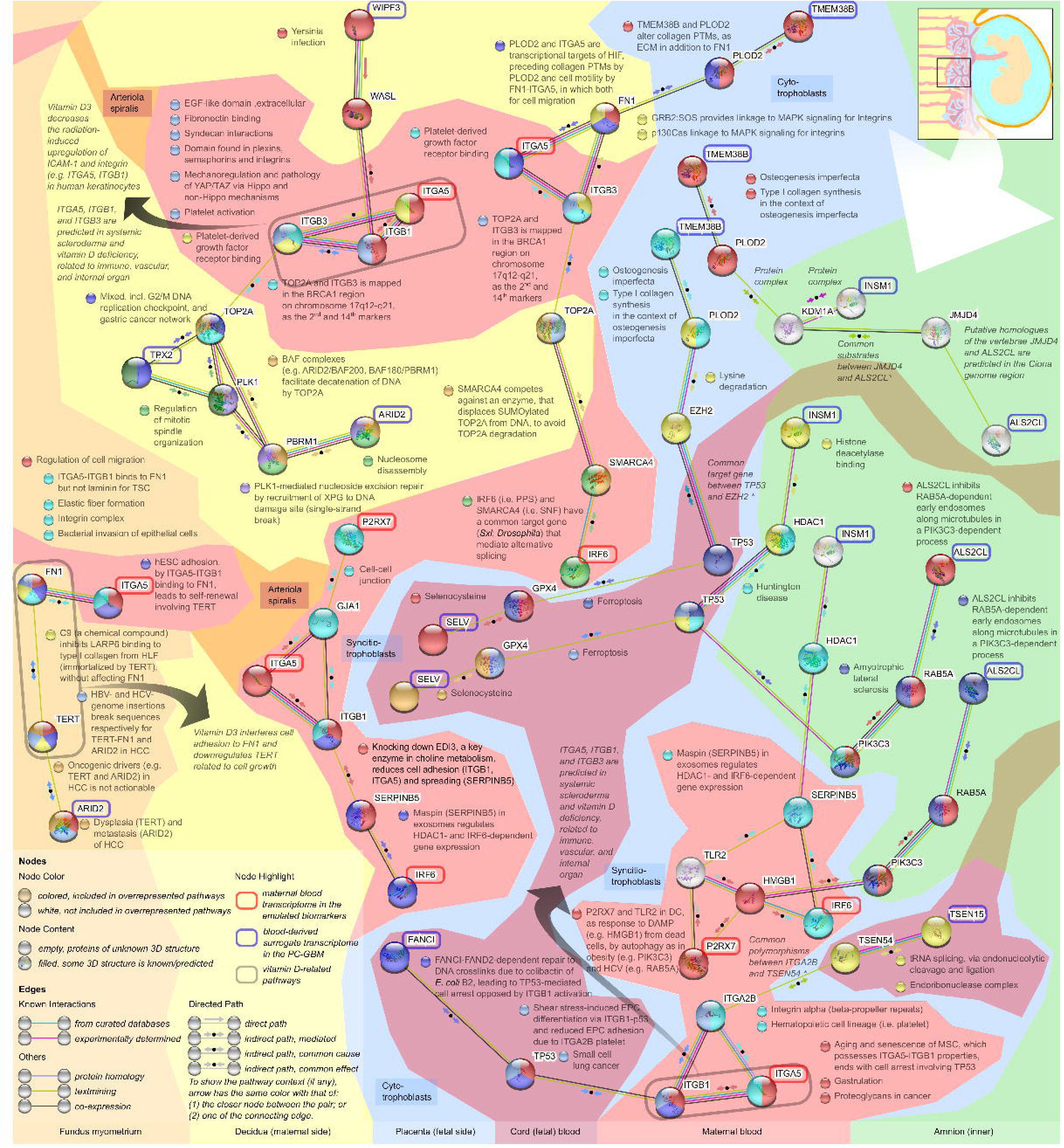
Networks and pathways in the context of the maternal-fetal interface. We used proteins in the shortest paths connecting all of the input pairs (biomarkers and the surrogate transcriptome as indicated by colored-highlighted names). Nodes represent proteins, for which the same colors of the nearest nodes indicate the same overrepresented pathway. The pathway descriptors are adjacent to the nodes in the same colors. The edges indicate both functional and physical protein associations with the directed paths [85]. The edge color indicates the type of interaction evidence. Proteins that overrepresented vitamin D-related pathways are surrounded by gray-colored highlights, with pointers to the descriptors. The colors of the areas indicate the tissue context. *, edge information instead of the pathway in the STRING database.

#### 3.5.1 Individual mRNAs, micro (mi)RNAs, post-translational modifications (PTMs), and biological effects

From the GeneCards human gene database, we retrieved gene information of genes under physiological conditions (Tables A.1, B.1, and B.2). We depicted tissue-specific protein expression based on gene information (Figure 4). Some of the proteins might not be expressed due to one to 39 miRNAs physiologically targeting the genes, not to mention different miRNA expressions under pathological conditions. From the DIANA miRNA tissue expression database, we queried all of the miRNAs in multiple tissues using comparisons between physiological and pathological conditions (Tables B.3 and B.4). The latter pathological conditions included preeclampsia with and without fetal growth restriction, and preterm delivery. However, except for SELV and INSM1, all other miRNA data were available but only for those in the placenta under preeclampsia with or without fetal growth restriction. We conducted a reanalysis by computing ORs of pathological conditions for every increase of 1 unit of log_2_[reads per million (RPM)] (Table A.2). miRNAs are depicted (Figure 4) if they targeted genes whose proteins of which fulfilled any of these criteria: (1) physiologically expressed in the placenta but the majority of miRNAs were not differentially expressed under any of the pathological conditions compared to the physiological condition (*ITGA5*, *P2RX7*, *IRF6*, *TSEN15*, and *ARID2*) or (2) physiologically not expressed in the placenta but the majority of miRNAs were differentially expressed under any of the pathological conditions compared to the physiological condition (*FANCI*, *TPX2*, *WIPF3*, and *TMEM38B*). For the latter, only miRNAs targeting *FANCI* and *TMEM38B* were significantly upregulated (Table A.2); thus, proteins were expressed in the placenta under preeclampsia for *TMEM38B* but only under preeclampsia with FGR for *FANCI* (Figure 4).

Information on PTM was also retrieved (Figure 4; Table B.5). We also considered a mapping between PTMs and biological implications (Table B.6) [36]. Protein overexpression in tissues of a pathological condition was inferred according to change in miRNAs, adding up those in a physiological condition (Figures 4 and A.2). These were: (1) *ITGA5* in blood, the uterus, and placenta; (2) *P2RX7* in blood and the placenta; (3) *IRF6* in the placenta; (4) *FANCI* in blood and the placenta; (5) *TSEN15* in blood; (6) *ARID2* in the placenta; and (7) *TMEM38B* in the placenta. All of the overexpressed proteins in tissues were modified by phosphorylation which was related to protein-protein interactions (PPIs), protein trafficking, cell-cycle division, and immune responses (Figure A.2). These proteins, except for P2RX7 and TSEN15, were also modified by ubiquitination which is related to protein stability, cell-cycle division, and immune responses. The ITGA5 and P2RX7 proteins were modified by glycosylation which is related to protein stability, PPIs, protein trafficking, protein thermodynamics and kinetics, and protein activity. Meanwhile, the P2RX7, FANCI and ARID2 proteins were modified by acetylation which is related to apoptosis, protein stability, transcription, and DNA repair. In addition to phosphorylation, glycosylation, and acetylation, the P2RX7 protein was also modified by (1) ribosylation which is related to apoptosis, cell signaling, transcription, and DNA repair and (2) palmitoylation which is related to protein membrane, protein trafficking, and cell signaling.

### 3.5.2 Protein-protein functional association network and pathway enrichment analysis

From the STRING functional protein association network, we retrieved a protein-protein functional interaction network such that all of the biomarker proteins (ITGA5, P2RX7, and IRF6) and those of the corresponding surrogate transcriptome were completely connected (Table A.3). The minimum score of the interaction was 0.4 (default setting). We set a maximum of 50 interactors for each of the first- and second-shell interactions with our input proteins. After maximum numbers were achieved, four proteins remained disconnected, which were encoded by the surrogate transcriptome in fetal tissues in the PC-GBM model: (1) INSM1 and ALS2CL in the amnion; (2) SELV in cord blood; and (3) TMEM38B in the placenta. But, if we queried only these proteins in the database, all of them were completely connected after a maximum of 50 and 20 interactors, respectively, at the first- and second-shell interactions. In the PC-GBM model, all of the biomarker transcripts in maternal blood corresponded to surrogate transcripts which were translated to INSM1, ALS2CL, SELV, and TMEM38B in the isolated network. Since maternal tissues (blood and the decidua) only interface with the placenta among fetal tissues in the isolated network, we focused on the corresponding surrogate transcript in the placenta, which was TMEM38B, to identify its interactors that openly connected to other surrogates in the isolated network. The only one that fit the criterion was PLOD2. This protein was then included to query all ITGA5 and IRF6 proteins in maternal blood, the transcripts of which corresponded to surrogates in the decidua and placenta, according to the PC-GBM model. The PLOD2 protein enabled connections between biomarkers and surrogate transcripts with those in the isolated network.

We also conducted a pathway enrichment analysis using all of the proteins and interactors based on multiple pathway databases which were integrated in the STRING platform. The number of interactors was reduced by including only those in the shortest paths connecting every pair of proteins in a single or a pair of tissues (Table A.4). This resulted in 37 proteins in total. The biomarkers and corresponding surrogate transcriptome in the PC-GBM encoded 32% (*n*=12 of 37) of the proteins. To interpret results of the pathway enrichment analysis, we selected significantly overrepresented pathways either for a single or a pair of tissues (Tables A.4 and B.7) by these criteria, according to a previous protocol [37]: (1) pathways, for each combination of genes, that had the number of background genes in this order of priority, i.e., 15 to 200, 10 to 14, and 201 to 500, or <10 and >500; and (2) pathways, for each combination and each database, that had both the highest strength of overrepresentation and the largest number of observed genes (allowing ties). Since pathway titles from PubMed are not always informative, we identified descriptors that were briefly informative and contextually relevant to our genes of interest, that were overrepresented in the pathways (Figure 5, Table B.7). The same criteria were also applied to filter only pathways related to vitamin D. Eventually, we determined a directed path in each edge based on pathways in the elaborated illustration (Figure 5) for the network and pathways in the context of the maternal-fetal tissue interface. If no common pathway was found, which included a pair of nodes connected by an edge, then we determined the directed path based on the edge information collected in the STRING database.

Eventually, based on either the pathway or edge information, we manually curated directed paths accompanying the edges (Figure 5). This is important for systematically interpreting the meaning of the edges. Most of the directed paths were indirect by common effect. The interpretation of path implications is elaborated (see Subsections 4.2.1 to 4.2.5).

## 4. Discussion

### 4.1 Summary of findings

We identified *ITGA5*, *IRF6*, and *P2RX7* as potential blood biomarkers to predict any-onset preeclampsia but not COVID-19 infection. These biomarkers represent the surrogate transcriptome of maternal-fetal interface tissues and were well-replicated to predict preeclampsia using a dataset without a vitamin D intervention. The ITGA5, IRF6, and P2RX7 transcripts had weights within the top two of any tissues in the PC-GBM model, and we subsequently selected from transcripts with the top three weights in each of the tissues (see Subsection 3.4). Without considering potential false positives due to COVID-19 infection, only the ITGA5 and IRF6 transcripts were needed to predict any-onset preeclampsia.

We found these predictors using the blood-derived surrogate transcriptome of maternal-fetal interface tissues for predicting preeclampsia but not COVID-19 infection. This method discovered these predictors not simply by chance, and identified the largest number of predictors among other methods, which were standard pipelines of predictor discovery using transcriptomic data, and those based on a previous preeclampsia gene set [30]. Predictions of the discovered biomarkers were mostly shared between preeclampsia and COVID-19 infection, including those from the previous preeclampsia gene set [30]. Since it was discovered during the pandemic but validated antecedently, the shared prediction was more likely because of the method of predictor discovery that was limited to identifying a blood biomarker of a condition, which should be unique among those conditions that shared endothelial dysfunction as a key pathophysiological derangement.

Post-analysis justification for the biological relevance of ITGA5, IRF6, and P2RX7 identified relationships between these blood biomarkers with the surrogate transcriptome, i.e., (1) FANCI, SELV, and TSEN15 in cord blood; (2) TPX2, WIPF3, and ARID2 in the decidua; (3) ARID2 in the fundus myometrium; (4) INSM1, ALS2CL, and TMEM38B in the amnion; and (5) TMEM38B in the placenta. These were justified at the levels of genes, miRNAs, PTMs, PPIs, enriched pathways, and directed paths. Both low- and high-level information implied the biomarker mechanism for predicting preeclampsia, the pathophysiological derangement related to polymicrobial infection and viral co-infection, the shared prediction with COVID-19 infection, and the non-replicability of the prediction under a vitamin D intervention.

### 4.2 Elaboration of the biomarkers and the biological relevance

To elaborate results of this study, we should consider the modeling pipelines. The surrogate transcriptome in maternal-fetal interface tissues was derived by the maternal blood transcriptome under non-preeclamptic conditions with or without PROM and terminated either preterm or at term. But, we developed the PC-GBM to identify important predictors of any-onset preeclampsia in maternal blood, that represented the transcriptome in maternal-fetal interface tissues. Among the transcripts involved in the pathophysiological derangement of any-onset preeclampsia in those tissues, the model only captured ones that were connected to the transcriptome in maternal blood under non-preeclamptic conditions. However, some of the connections changed due to preeclampsia; thus, the model did not take those into account. The model was also chosen only if it was well-replicated when using a dataset with no preventive intervention, but not those with a vitamin D intervention. Any biological process that was interfered with by vitamin D would impair the performance of a model that was not significantly predictive (i.e., an AUROC interval of <0.5). Eventually, the final biomarkers were those that fulfilled the criteria to identify a few, well-replicated predictors of any-onset preeclampsia but not COVID-19 infection. But, we needed to differentiate which results of the biomarker predictions were likely due to any-onset preeclampsia or COVID-19 infection using a decision tree.

#### 4.2.1 Polymicrobial infection of fetal tissues in preeclampsia implied by ITGA5

The majority of the predicted events (73.70%) among any-onset preeclampsia samples in the development dataset were solely defined by the ITGA5 transcript with a Z-score of ≥1.1 (Figure 4). This included only a minority of the predicted events (6.34%) among those with COVID-19 infection. Expression of the ITGA5 transcript, as shown by the PC-GBM, was inverse (negatively weighted) to those of FANCI, SELV, and TSEN15 in cord blood.

A pathway from PubMed [38], overrepresented by the genes including *FANCI* (Table B.7), indicated a repair response to DNA damage by crosslinks due to colibactin of *Escherichia coli* B2. This protein had a common-cause, indirect path to p53 that mediates cell arrest (Figure 5). This might be a DNA-repair response of syncitiotrophoblasts of the placenta, of which either a transcript-containing exosome or protein of FANCI might be secreted into cord blood. This was supported by (1) the transcript being overexpressed in cord blood of the derivation dataset; (2) most of the miRNAs targeting FANCI being downregulated under preeclampsia with FGR (Figure 4; Table A.2); (3) the protein also being overexpressed in blood plasma (Figure 4; Table B.2); and (4) the PTMs including protein trafficking and PPIs as implications (Figure A.2). Expression of the FANCI transcript in cord blood was inversely weighted with the ITGA5 transcript in defining predicted preeclampsia, according to the PC-GBM (Figure 4). The DNA-repair response, including the cell-arrest mechanism, might be impaired in syncitiotrophoblasts which mediate protein trafficking and PPIs between maternal and fetal blood. In addition, the role of FANCI in syncitiotrophoblasts of the placenta was also supported by its protein overexpression in this tissue due to downregulation of miRNAs under preeclampsia with FGR (Figure 4; Table A.2). However, the PC-GBM model did not show that ITGA5 corresponded to FANCI in the placenta, because the surrogate transcriptome model was derived under non-preeclamptic conditions.

Both ITGA5 and FANCI were connected via ITGB1-p53 in the shortest path (Figure 5). Epithelial progenitor cell (EPC) differentiation (Figure 5), which is probably applied to placental trophoblasts, is induced by shear stress via by the ITGB1-p53 pathway, leading to differentiation into syncitiotropblasts at the placenta-maternal blood interface [39]. ITGB1 activation opposes the p53-mediated cell arrest [38]. But, at the placenta-decidua interface, a membrane-bound ITGB1-ITGA5 receptor may induce cell arrest as inferred from that in mesenchymal stromal cells (MSCs) [40], instead of opposing p54-mediated cell arrest. This is probably because placental trophoblasts undergo the epithelial-to-mesenchymal transition (EMT) [41], which changes the cell arrest response related to ITGB1 at the placenta-decidua interface. Meanwhile, EPC adhesion is reduced due to ITGA2B platelets [39]. ITGA2B also forms a complex with ITGA5 as a membrane-bound receptor of platelets, genes of which were overrepresented in the SMART (SM00191), KEGG (hsa04640 and hsa05205), and GO Process (GO:0007369) pathways (Table B.7).

Common polymorphisms exist between ITGA2B and TSEN54 [42], of which the latter forms an endonuclease complex with TSEN15 (Figure 5). The *TSEN54* and *TSEN15* genes were overrepresented in the GO Process (GO:0006388) and COMPARTMENTS (GOCC:1902555) pathways (Table B.7). The surrogate transcript of TSEN15 in the PC-GBM was also inversely weighted with the ITGA5 transcript in defining predicted preeclampsia. Since the absence of polymorphisms is most likely in any pregnant women, preeclampsia prediction using the ITGA5 transcript may be replicable only in this situation.

The third surrogate transcript of cord blood in the PC-GBM was SELV, which inversely corresponded to ITGA5 (Figure 4). The Selv protein can upregulate Gpx4 transcript expression (Figure 5) in the liver and testes under a specific amount of dietary selenium given to mice [43]. A meta-analysis of eight observational studies showed lower selenium concentrations of either maternal or cord blood from Asian preeclamptic women (mean difference -9.77, 95% CI -16.76 to -2.79; *n*=299; *I^2^* 92%), while a meta-analysis of three randomized-controlled trials showed selenium supplementation reduced the relative risk of preeclampsia (0.28, 95% CI 0.08 to 0.84; *n*=218; *I^2^*0%) [44]. The *GPX4* and *TP53* genes were overrepresented in the WikiPathways (WP4313) and KEGG (hsa04216) pathways (Table B.7). Both of these pathways are ferroptosis. This may indicate an impaired intracellular antioxidant system due to either ITGA5-related p53 upregulation or SELV-related GPX4 downregulation, which may share a common cause. Ferroptosis, including GPX4, is also involved in bacterial infections and polymicrobial sepsis [45], including by *E. coli* [46]. This provides a potential link to the DNA-repair response involving FANCI.

Nevertheless, mRNA expression of SELV was only found in testes but with an unknown location for protein overexpression under physiological conditions (Tables B.1 and B.2). Unlike the FANCI transcript that was overexpressed in the placenta with protein overexpression in blood plasma, secretion from syncitiotrophoblasts of the placenta to cord blood is unclear for SELV either as a transcript-containing exosome or protein. Meanwhile, the shortest paths connecting ITGA5, FANCI, SELV, and TSEN15 commonly involved p53 (Figure 5), in which PPIs were likely intracellular within a common cell type. A potential cell type is circulating trophoblasts; however, previous studies only investigated this cell type in maternal blood up to 4 weeks postpartum [47, 48], but not in cord (fetal) blood. Circulating trophoblasts were those from extravillous trophoblasts [47], which also require the EMT, as occurs in the placenta-decidua interface. Future investigations need to identify circulating trophoblasts in cord blood. This may help elucidate pathophysiological derangement of preeclampsia, that involves downregulation of FANCI, SELV, and TSEN15 in cord blood, corresponding to upregulated ITGA5 in maternal blood.

#### 4.2.2 Polymicrobial infection of uterine tissues in preeclampsia implied by ITGA5

Unlike the surrogate transcriptome in cord blood as shown by the PC-GBM (Figure 4), expression of the ITGA5 transcript was alike (positively weighted) to those of TPX2, WIPF3, and ARID2 in the decidua, but inverse (negatively weighted) to those of ARID2 in the myometrium. No proteins of these surrogate transcripts were found to be overexpressed in the uterus under a non-preeclamptic condition (Table B.2). However, we did not find miRNA data in the uterus that could justify the absence or presence of protein expressions in this tissue, as in the placenta (Figure 4); thus, the proteins may also be overexpressed in the uterus under some circumstances. All of the genes in the shortest path connecting ITGA5 and WIPF3 were overrepresented in a KEGG (hsa05135) pathway (Table B.7). This indicated that ITGA5-ITGB1 and WIPF3 have a synergistic, common effect (Figure 5), which was that on WASL, resulting anti-phagocytosis and disruption of the actin cytoskeleton, according to the KEGG pathway. It describes Yersinia infections, or more generally, virulence of many gram-negative bacteria by the type III secretion system for injecting toxins to immune and epithelial cells, leading to immune evasion and/or cell invasion, which are location-specific via binding of the bacterial adhesins to ITGA5-ITGB1 [49].

Other surrogate transcripts of the decidua in the PC-GBM were connected to ITGA5 via ITGB3 (Figure 5). Both ITGA5 and ITGB3 were overrepresented in a GO Function (GO:0005161) pathway (Table B.7) for platelet-derived growth factor receptor binding. Hepatocyte growth factor, which is mostly derived from platelets, forms a complex among ITGA5-ITGB1, ITGAV-ITGB3, and Met [50]. Overrepresentation by ITGB1 and ITGB3 was also found in a KEGG (hsa04611) pathway for platelet activation and five pathways from five different databases (Table B.7). The KEGG pathway depicted collagen binding to ITGA2-ITGB1 of platelets, which leads to complement and coagulation cascades via ITGA2B-ITGB3. Therefore, upregulation of ITGA5 might not only result in anti-phagocytosis and disruption of the actin cytoskeleton in immune and epithelial cells in the decidua, but also platelet adhesion with the complement and coagulation cascades.

A pathway overrepresentation by ITGB3 and TOP2A connected ITGA5 to other surrogate transcripts of the decidua in the PC-GBM model (Figure 5). This pathway was retrieved by STRING from PubMed, which mapped the *TOP2A* and *ITGB3* genes in the BRCA1 region on chromosome 17q12-q21 (Table B.7). These genes probably share a common cause affecting the BRCA1 region such that both of the genes are upregulated under a preeclamptic condition. Two pathways from STRING clusters and PubMed were overrepresented by TOP2A with respect to TPX2 and ARID2, which were surrogate transcripts of the decidua in the PC-GBM model (Table B.7). These are related to DNA replication and decatenation respectively involving TOP2A-TPX2 and TOP2A-ARID2-PBRM1. The common cause affecting the BRCA1 region may lead to cell proliferation in the decidua. This is probably a response to protect uterine blood vessels from infection-related endothelial dysfunction, because MSCs in the decidua, particularly extracellular vesicles, exhibited increased proliferation and attachment of endothelial cells *in vitro* (i.e., human umbilical vascular endothelial cells, HUVECs) treated with either bacterial lipopolysaccharide or serum from preeclamptic women [51].

To this point, the biological relevance of ITGA5 prediction for terminal branch A (Figure 4) implied “normal” placentation at placenta-maternal blood and placenta-decidua interfaces, but involving polymicrobial infection and platelet-related responses. A previous study identified clinically relevant subclasses of preeclampsia [52]: (1) a precondition to other subclasses, which is healthy placenta consisting of maternal non-preeclamptic term delivery, non-infection preterm delivery, and maternal preeclampsia which was mostly similar to LOPE; (2) canonical preeclampsia, mostly similar to EOPE; (3) immunological preeclampsia, mostly similar to FGR with or without preeclampsia; (4) infection-related preterm delivery; and (5) any other subclasses with chromosomal abnormalities. However, this contradicts the revised two-stage model that proposed maternal preeclampsia in the first subclass having an abnormal placenta, in which placentation is normal but undergoes uteroplacental malperfusion at term [2]. Herein, we proposed that malperfusion at term would have not been manifested to LOPE, had it not been preceded by adequate placental response to hematogenous infection. An infection was implied by terminal branch A which mostly included the predicted events among either EOPE or LOPE samples in the development dataset, consistent with the first subclass as proposed by a previous study [52]. Although the fourth subclass was also related to infections, it was mostly chorioamnionitis, for which cases were likely because of an ascending, genital infection instead of that from a hematogenous route [53].

The hallmark of abnormal placentation in EOPE is failure of physiological spiral artery remodeling at the myometrium-decidua interface [54]. Meanwhile, terminal branch A implied “normal” placentation, which also included the majority of the predicted events among EOPE samples in the development dataset. Yet, unlike the surrogate transcript of ARID2 in the decidua, the ITGA5 transcript was inverse (negatively weighted) to that of ARID2 in the fundus myometrium, as shown by the PC-GBM model (Figure 4). In the context of this tissue, the ITGA5 protein was connected to ARID2 by the shortest path including FN1 and TERT (Figure 5). All four genes were overrepresented in a GO Process (GO:0030334) pathway for regulating cell migration, while three of the genes, excluding ITGA5, were overrepresented in a PubMed pathway (Table B.7). The latter pathway included a description of hepatitis C virus genome insertion that breaks sequences of ARID2 [55]. This may explain the downregulation of ARID2, in which the transcript expression is inverse to that of ITGA5 in the PC-GBM model. Since co-infection with bacteria and viruses reasonably has a lower probability compared to that of only a bacterial infection, this is coincidentally consistent with the lower weight of ITGA5 to the surrogate ARID2 transcript of the fundus myometrium in the PC-GBM model. In this model, the top 20 weights in the maternal blood transcriptome within the fundus myometrium were not within the top five in any tissues. Instead, the weight of ITGA5 corresponding to ARID2 in the fundus myometrium was the top 1,974^th^ in the PC-GBM model (Figure 4).

#### 4.2.3 Viral co-infection in early-onset preeclampsia implied by ITGA5-IRF6

The remaining predicted events (26.30%) among any-onset preeclampsia samples in the development dataset were defined by the ITGA5 transcript with a Z-score of <1.1 (Figure 4). Contrary to the previously described effect, downregulation of ITGA5 may result in reduced differentiation of placental trophoblasts into syncitiotrophoblasts, which normally occurs at the placenta-maternal blood interface. Accordingly, p53-mediated cell arrest is also reduced at the placenta-decidua interface. This may lead to typical placentation in EOPE. In the decidua, downregulation of ITGA5 was also followed by the surrogate transcriptome, according to the PC-GBM model. This may lead to reduced proliferation of MSCs in the decidua, followed by reduced proliferation and attachment of endothelial cells. However, since the ITGA5 transcript with a Z-score of ≥1.1 was the majority of the predicted events among either EOPE or LOPE samples, the IRF6 transcript with a Z-score of ≥-0.73 was needed to define the predicted events of preeclampsia, especially EOPE.

Terminal branches B, C, and D (Figure 4) indicated relative upregulation of the IRF6 transcript, but the cutoff value was higher for terminal branches B and C compared to that of terminal branch D (Z-scores of -0.73 vs. -0.81). If only samples with either EOPE or isolated FGR were used in the development dataset, the proportions of the predicted events or nonevents were shifted toward the terminal branches B/C or D, respectively. Among the emulated biomarkers, only the IRF6 transcript was connected to the surrogate transcriptome in the placenta with or without preeclampsia. This probably explains the similarity of placental characteristics of EOPE and isolated FGR.

Upregulation of *IRF6* may result from a reduction in negative feedback to IRF6 transcription. The protein has a synergistic effect with *SERPINB5* (Figure 5). Both genes and HDAC1 were overrepresented in a PubMed pathway (Table B.7) which indicated such an effect [56]. Meanwhile, the genes of *ITGA5-ITGB1* and *SERPINB5* genes were overrepresented in another PubMed pathway that showed a common cause of a reduction in cell adhesion (*ITGA5-ITGB1*) and spreading (*SERPINB5*) [57]. Since ITGA5 was downregulated in terminal branches B to E (Figure 4), *SERPINB5* expression was likely downregulated. According to that pathway [57], since *IRF6*-dependent gene expression is regulated by SERPINB5 a reduction of which avoids the downstream effect of IRF6, then transcript expression is upregulated if the downstream effect provides negative feedback to IRF6 transcription. The interferon regulatory factor (IRF) family is important for inducting interferons in both antiviral and antimicrobial responses, particularly IRF6 with a downstream effect on transcription of type II interferon [58]. This interferon provides negative feedback to its transcription via interleukin (IL)-10 [59, 60]. Avoiding the downstream effect of IRF6 may cancel the negative feedback; thus, IRF6 upregulation is maintained without its antiviral and antimicrobial effects.

Without the protective mechanism, damage-associated molecular patterns (DAMPs) may be identified. DAMPs include HMGB1 in the shortest path from IRF6 to ALS2CL (Figure 5) which was the surrogate transcriptome of the amnion in the PC-GBM (Figure 4). The HMGB1 protein was proposed to be one of the effectors of sterile inflammation that may cause preeclampsia, in which one of the mediating inflammasomes (i.e. pyrin) can lead to inactivation of Rho GTPases and microtubule disruption due to microbial infection (i.e., pathogen-associated molecular patterns, PAMPs) but not DAMPs [61]. This may be related to a PubMed pathway overrepresented by ALS2CL and two interactors (viz., RAB5A and PIK3C3) in its shortest path to IRF6 (Figure 5; Table B.7). The pathway describes the role of ALS2CL in the inactivation of Rho GTPases and microtubule disruption [62]. However, the surrogate ALS2CL transcript of the amnion was inverse (negatively weighted) to that of IRF6, which may indicate loss of response following the pyrin inflammasome, regardless of the triggers, either PAMPs or DAMPs. Nonetheless, the IRF6 transcript would have not upregulated, had it no microbial infection; thus, sterile inflammation might not cause preeclampsia.

Since the HDAC1 protein also has a synergistic effect with SERPINB5 (Figure 5), the downstream effect of this protein was also avoided as was that of IRF6. A pathway of GO function (GO:0042826) was overrepresented by HDAC1, TP53, and INSM1. The pathway describes histone deacetylase (HDAC) binding; thus, avoiding the downstream effect of HDAC1 would result in HDAC inhibition. This was demonstrated to result in (1) a dose-dependent increase of chymase expression in HUVECs, the upregulation of which was found in the maternal endothelium under preeclampsia and (2) generation of chymase-dependent angiotensin II, as reported in several cardiovascular diseases [63]. In terminal branch A (Figure 4), we indicate ITGA5-related p53 upregulation or SELV-related GPX4 downregulation. If the regulation is inverted in the other terminal branches of the decision tree (Figure 4), then p53 downregulation is consistent in avoiding HDAC binding. Regulation of p53 connects all of the surrogate transcripts of fetal tissues in the PC-GBM model, that were derived by all the emulated biomarkers (Figures 4 and 5).

The surrogate TMEM38B transcript was both inverse and alike (negatively and positively weighted) to that of IRF6, in which the latter weight made IRF6 a lesser rank of biomarkers in deriving the surrogate transcriptome of the placenta. This implied the surrogate TMEM38B transcript of the placenta is affected by the presence or absence of other substances in the same tissues such that the expression of TMEM38B is alike to that of IRF6 under preeclampsia, but the expression is inverse in other tissues if such substances are absent or present under non-preeclamptic conditions. This may be related to EZH2 which only exists if we identified the shortest paths between TMEM38B and either the emulated biomarkers or the surrogate transcriptome in the context of the cord blood-placenta interface (Figure 5). The shortest paths did not include EZH2 if the paths were identified in the context of placenta-amnion and decidua-placenta interfaces. A KEGG (hsa00310) pathway was overrepresented by EZH2 and PLOD2 (Table B.7). Both EZH2 and PLOD2 are involved in lysine degradation respectively resulting in carnitine-glycine and protein 5-galactosyloxylysine as either end or side products. We could find no evidence for an association of the latter with preeclampsia, but a systematic review identified carnitine-related metabolites and glycine as metabolomics associated with preeclampsia [64]. Notably, acyl carnitine and glycine were significantly higher in preeclamptic women and cord blood, respectively, compared to normotensive controls and maternal blood; however, these did not individually predict preeclampsia [65]. This is probably because the role of EZH2 may be minor if it is related to expression of the TMEM38B transcript that is alike to IRF6. In this circumstance, the absolute weight of the IRF6 transcript to the surrogate transcript of TMEM38B was less than the inverted one (Figure 4).

The surrogate TMEM38B transcript of the amnion in the PC-GBM was inverse (negatively weighted) to IRF6 (Figure 4). In addition to the shortest path that included EZH2, we also identified other paths that connected IRF6 in maternal blood and TMEM38B in the context of the placenta-amnion and decidua-placenta interfaces (Figure 5). All of the shortest paths including TMEM38B required PLOD2. Both were overrepresented in UniProt Keywords (KW-1065) and WikiPathways (WP4786) pathways (Table B.7). If TMEM38B transcript was downregulated, then the possible downstream effect would be impaired type I collagen synthesis, particularly in the context of the placenta-amnion. In this circumstance, since a protein complex was formed by PLOD2, KDM1A, and INSM1 (Figure 5), downregulation of the latter transcript would result in the same downstream effect with downregulation of the TMEM38B transcript, as implied by the inverted weights of those surrogates to the emulated biomarkers and the PC-GBM model (Figure 4). Even if a pregnant woman was predicted to be in terminal branch C (Figure 4), in which downregulation of P2RX7 would correspond to upregulation of INSM1, the downstream effect still follows downregulated TMEM38B with upregulated IRF6. Therefore, this probably impairs type I collagen synthesis in the amnion. However, we found no miRNA data in the amnion that could justify possible protein expression of TMEM38B in this tissue, as that in the placenta (Figure 4); thus, the impaired synthesis in the amnion might also never occur.

Terminal branches B, C, and D were all defined by downregulated ITGA5 and upregulated IRF6, which corresponded to upregulation of the TMEM38B protein in the placenta, according to the biomarkers, the PC-GBM model, and the miRNAs (Figure 4). In the context of the decidua-placenta interface (Figure 5), the shortest path also included PLOD2, FN1, ITGB3, TOP2A, and SMARCA4. In contrast to the effect, as described previously, downregulation of ITGA5 may result in reduced cell proliferation in the decidua, leading to an impaired protective response for uterine blood vessels against infection-related endothelial dysfunction [51]. Viral co-infection may have a putative role in this circumstance. Furthermore, impaired trophoblasts may also be related to the shortest path between ITGA5 and TMEM38B via FN1 and PLOD2 (Figure 5). Transcripts of ITGA5, FN1, and PLOD2 were overrepresented in a PubMed pathway (Table B.7) which describes regulation of cell migration by hypoxia via collagen PTMs (PLOD2) and cell motility (FN1-ITGA5) [66]. Meanwhile, transcripts of FN1, PLOD2, and TMEM38B were overrepresented in a PubMed pathway which describes involvement of TMEM38B in collagen PTMs by PLOD2 in order to alter the extracellular matrix (ECM) in addition to FN1 [67].

Downregulated ITGA5 in terminal branches B, C, and D would only correspond to upregulated TMEM3B in the placenta if the IRF6 transcript is upregulated (Figure 4). While the shortest path exists between IRF6 and TMEM38B, a possible explanation is not straightforward by the interactors along the path. This is probably because the STRING interaction is single-species. This means a PPI only semantically involves a protein name from another species, i.e., Sxl of *Drosophila*, but did not provide an alternative interaction by the human homologue. The *IRF6*-*SMARCA4* and *SMARCA4*-*TOP2A* genes were overrepresented in two PubMed pathways (Table B.7). The pathways respectively describe how (1) the IRF6 and SMARCA4 proteins have a common target gene (*Sxl*) [68], a *Drosophila* homologue of human antigen R (currently ELAVL1) which stabilizes AU-rich RNA element (ARE)-containing mRNAs [69] and (2) the SMARCA4 protein avoids TOP2A degradation [70]. A higher level of the IRF6 protein may allow more SMARCA4 proteins to avoid TOP2A degradation during cell proliferation in the decidua, which is nevertheless reduced, as inferred from the downregulation of ITGA5. However, human antigen R was identified in placental homogenates of preeclamptic women, which induced aggregation of cytoplasmic stress granules in a human trophoblast cell line (HTR-8/SVneo cells) [71]. Upregulated IRF6 in terminal branches B, C, and D (Figure 4) may be transcriptionally involved in the pathogeneses of preeclampsia and FGR, via human antigen R. This may explain the shared pathophysiological derangement of both conditions related to impaired trophoblasts which are important for spiral artery remodeling during placentation [72]. Upregulated IRF6 in maternal blood may be a part of exosomes which undergo endocytosis into cells in the placenta and was translated into the protein (Figure 4), which transcriptionally activates human antigen R. If it is inhibited in human lung fibroblasts by transfecting cells with its small interfering (si)RNA, a significant reduction in FN1 also occurred [73]. This implied that the upregulation of IRF6 increases FN1 as negative feedback to impaired, ITGA5-mediated cell migration. Yet, this physiological protection is inadequate in conditions defined by terminal branches B, C, and D (Figure 4). Alternatively, there is probably another substance which behaves similarly to the siRNA of human antigen R, leading to the reduction of FN1 [73].

#### 4.2.4 Shared predictions between preeclampsia and COVID-19 by ITGA5-IRF6-P2RX7

As described previously, a PubMed pathway was overrepresented by RAB5A, PIK3C3, and the surrogate ALS2CL transcript of the amnion in its shortest path to IRF6, which was inversely connected (negatively weight) (Figures 4 and 5). The RAB5A and PIK3C3 transcripts were also overrepresented in a PubMed pathway with HMGB1 and P2RX7 (Table B.7). The pathway describes P2RX7 and TLR2 of dendritic cells (DCs) as a response to HMGB1 by autophagy, similar to those in obesity and hepatitis C virus infection respectively involving PIK3C3 and RAB5A [74]. Terminal branch B (Figure 4) defined the predicted events of both preeclampsia and COVID-19 infection, in which the IRF6 and P2RX7 transcripts in maternal blood were upregulated (with respective Z-scores of ≥-0.73 and ≥0.13). This implied an increasing response to DAMPs.

Furthermore, the P2RX7 transcript had the shortest path to the surrogate INSM1 transcript of the amnion in the PC-GBM model, in both those related and unrelated to IRF6 (Figure 5). These respective paths were either P2RX7-TLR2-SERPINB5-HDAC1-INSM1 or P2RX7-HMGB1-PIK3C3-TP53-HDAC1-INSM1. The surrogate INSM1 transcript of the amnion in the PC-GBM model was inverse (negatively weighted) to P2RX7 (Figure 4). This is consistent with the previous description of avoiding the downstream effect of HDAC1. HDAC inhibition results in chymase-dependent angiotensin II generation in preeclampsia [63]. This was also proposed for COVID-19, which is a major non-renin, non-angiotensin-converting-enzyme (ACE) blood pressure regulatory system that activates transforming growth factor (TGF)-β, matrix metalloproteinase (MMP)-9, and thrombin-plasmin, which are respectively related to structural injury, organ remodeling, and enhanced coagulation [75]. In addition to chymase-dependent angiotensin II, TGF-β [76], MMP-9 [77], and thrombin-plasmin [78] were also proposed to have roles in for preeclampsia. Chymase-dependent angiotensin II may also be a therapeutic target for preeclampsia, including one that is superimposed by COVID-19 infection [79].

#### 4.2.5 Non-replicability of the prediction in datasets with a vitamin D intervention by ITGA5

Three PubMed pathways related to vitamin D were overrepresented by FN-TERT [80], ITGA5-ITGB1 [81, 82], and ITGA5-ITGB1-ITGB3 (Table B.7) [82]. Vitamin D3 interferes with cell adhesion to FN1 and downregulates TERT related to cell growth [80], and decreases radiation-induced upregulation of ITGA5-ITGB1 [81], which is related to immune, vascular, and internal organs in vitamin D deficiency [82]. All of the vitamin D-related pathways were identified in the shortest paths that connected ITGA5 to surrogate transcripts in the PC-GBM model (Figures 4 and 5). Vitamin D supplementation was associated with a reduced risk of preeclampsia based on 27 randomized-controlled trials (OR 0.37, 95% CI 0.26 to 0.52; *I^2^* 0%) [83]. The effect of vitamin D on the risk of preeclampsia and the regulation of ITGA5 implied that placentation under the predicted events by terminal branch A (Figure 4) cannot be considered normal, as previously proposed [2]. Vitamin D probably ameliorates dysregulated placentation under polymicrobial infection and platelet-related responses, which may be classified as the first subclass of preeclampsia, as a precondition to other subclasses [52]. Because vitamin D may interfere with the process leading to the precondition subclass, the PC-GBM was not well-replicated in datasets with a vitamin D intervention.

### 4.3 Strengths and limitations

The proposed method of predictor discovery in this study identified blood transcripts that were not extremely-expressed genes, but these could predict preeclampsia but not COVID-19 infections, and were guided to derive transcripts in condition-specific tissues. This result could not be achieved by standard pipelines, although these pipelines used datasets before the pandemic. Conversely, the previous gene set [30] used in this study could not significantly discover eligible biomarkers, although genes were discovered by standard pipelines during the pandemic and also validated by a dataset before that time. Taken together, these findings demonstrated that the proposed method could discover predictors of a condition among others that shared common pathophysiological derangement in endothelial dysfunction.

However, there are several limitations of this study. Validation by RT-qPCR should be conducted for the proposed biomarkers and their surrogate genes. A larger sample size is needed to allow development of a more-accurate and more-generalized prediction model using these biomarkers. To avoid excessive costs, early predictions and low-cost preliminary predictions, e.g., utilizing electronic health records [11], would be preferred. The performance of the combined prediction should be validated, and its impact should be evaluated. Nevertheless, this study provided extensive screening of potential blood biomarkers that could predict preeclampsia but not COVID-19 infection which disrupted previously established biomarkers for preeclampsia [4, 5]. It is costly to experimentally screen many biomarkers, and it is also not scalable to identify biomarkers by only interpreting previous studies. Utilizing shared datasets and annotation databases, we could resolve those problems, particularly in such a way as to avoid false discoveries due to endothelial dysfunction.

## 5. Conclusions

A PC-GBM model using the blood-derived surrogate transcriptome could replicate the predictive performance in an independent dataset without an intervention unlike models with any algorithms using the blood transcriptome. The PC-GBM model could predict both early-and late-onset preeclampsia. From this model, we identified ITGA5, IRF6, and P2RX7 as potential blood biomarkers to predict preeclampsia but not COVID-19 infection, that represent the surrogate transcriptome of maternal-fetal interface tissues. By modeling the blood-derived surrogate transcriptome in target tissues, the proposed method significantly discovered eligible biomarkers, outperforming those found by a differential expression analysis and a previous gene set. Independent validation of the decision tree of potential biomarkers is needed using RT-qPCR analyses of maternal blood.

## Data Availability

All data produced are available online at:
https://www.ncbi.nlm.nih.gov/geo/query/acc.cgi?acc=GSE73685
https://www.ncbi.nlm.nih.gov/geo/query/acc.cgi?acc=GSE108497
https://www.ncbi.nlm.nih.gov/geo/query/acc.cgi?acc=GSE85307
https://www.ncbi.nlm.nih.gov/geo/query/acc.cgi?acc=GSE86200
https://www.ncbi.nlm.nih.gov/geo/query/acc.cgi?acc=GSE149437
https://www.ncbi.nlm.nih.gov/geo/query/acc.cgi?acc=GSE177477
https://www.ncbi.nlm.nih.gov/geo/query/acc.cgi?acc=GSE192902

https://github.com/herdiantrisufriyana/pest

## Appendices

Appendix A, Supplemental Notes.

Appendix B, Supplemental Spreadsheet.

## Author contributions

**H.S.:** Conceptualization; Data curation; Formal analysis; Funding acquisition; Investigation; Methodology; Project administration; Resources; Software; Visualization; Writing – original draft. **H.M.S.:** Data curation; Project administration; Writing – review & editing. **A.R.M.:** Project administration; Validation; Writing – original draft. **Y.W.W.:** Methodology; Software; Supervision; Writing – review & editing. **E.C.Y.S:** Conceptualization; Funding acquisition; Methodology; Project administration; Resources; Software; Supervision; Writing – review & editing. All authors approved the final version of the article.

## Code availability

The analysis codes are available at https://github.com/herdiantrisufriyana/pest.

## Data availability

The datasets are publicly available in the Gene Expression Omnibus (GEO) database.

## Acknowledgements

This study was funded by: (1) the Lembaga Penelitian dan Pengabdian kepada Masyarakat (LPPM) Universitas Nahdlatul Ulama Surabaya in Indonesia (grant no.: 161.5.1/UNUSA/Adm-LPPM/III/2021) to Herdiantri Sufriyana; (2) the Ministry of Science and Technology (MOST) in Taiwan (grant nos.: MOST109-2221-E-038-018 and MOST110-2628-E-038-001) to Emily Chia-Yu Su; and (3) the Higher Education Sprout Project from the Ministry of Education (MOE) in Taiwan (grant no.: DP2-110-21121-01-A-13) to Emily Chia-Yu Su. These funding bodies had no role in the study design; in the collection, analysis, and interpretation of the data; in the writing of the report; or in the decision to submit the article for publication.

## Footnotes

1 The publication erroneously reported 10 males of symptomatic COVID-19 individuals (Table 1), which was unmatched with the total number. We verified only 9 males of them in the dataset. No erratum was found.

2 While the publication reported this number in Table 1, which was matched with total numbers by sex and age, only 18 uninfected individuals were reported in the main text. No erratum was found.

