## Supplemental Notes for "Blood biomarkers representing maternal-fetal interface tissues used to predict early-and late-onset preeclampsia but not COVID-19 infection"

Table A.1 The biomarkers and the surrogate transcriptome under physiological condition.

| Gene symbol | Description | Location * | Gene information archived web page |
| --- | --- | --- | --- |
| ITGA5 | Integrin Subunit Alpha 5. A part of a heterodimeric integral membrane protein composed of this protein and the beta 1 subunit to form a fibronectin receptor. | Plasma membrane | [1] |
| P2RX7 | Purinergic Receptor P2X 7. A nuclear purinoceptor for ATP, which is a ligand-gated ion channel, coupled to changes in gene expression to form membrane pores permeable to large molecules in ATP-dependent lysis of antigen-presenting cells, and a scavenger receptor for engulfment of apoptotic cells in absence of ATP. | Plasma membrane | [2] |
| IRF6 | Interferon Regulatory Factor 6. A probable DNA-binding transcriptional activator that plays a role in epithelial cell proliferation by similarity to that in appropriate epidermal development. | Nucleus | [3] |
| FANCI | Fanconi Anemia Complementation Group I. Plays a role in DNA repair for maintenance of chromosomal stability. Specifically binds both single-stranded DNA and double-stranded DNA. | Nucleus | [4] |
| SELV (SELENOV) | Selenoprotein V. May be involved in a redox-related process. The UGA codon in selenoprotein mRNAs is recognized as a selenocysteine codon rather than as a stop signal. | Cytosol | [5] |
| TSEN15 | tRNA Splicing Endonuclease Subunit 15. A non-catalytic subunit of the endonuclease complex, that cleaves pre-tRNA to release the intron, and that establishes a link between pre-tRNA splicing and pre-mRNA 3’-end formation. | Nucleus | [6] |
| TPX2 | Targeting Protein for Xklp2 (TPX2) Microtubule Nucleation Factor. A spindle assembly factor during apoptosis in chromatin- and/or kinetochore-dependent microtubule nucleation. | Cytoskeleton | [7] |
| WIPF3 | Wiskott-Aldrich Syndrome (WAS)/WAS Like (WASL) Interacting Protein Family Member 3. May be a regulator of cytoskeletal organization. | Cytoskeleton | [8] |
| ARID2 | AT-Rich Interaction Domain 2. A DNA-binding protein containing ARID to facilitate ligand-dependent transcriptional activation and repression of different genes by chromatin remodeling. | Nucleus | [9] |
| INSM1 | Insulinoma-Associated Protein (INSM) Transcriptional Repressor 1. A sequence-specific DNA-binding transcriptional regulator to promote cell cycle signaling arrest and inhibition of cellular proliferation. | Nucleus | [10] |
| TMEM38B | Transmembrane Protein 38B. Monovalent cation channel required for maintenance of rapid intracellular calcium release. May act as a potassium counter-ion channel that functions in synchronization with calcium release from intracellular stores. | Endoplasmic reticulum | [11] |
| ALS2CL | ALS2 C-Terminal Like. Acts as a guanine nucleotide exchange factor (GEF) for Rab5 GTPase. Regulates the ALS2-mediated endosome dynamics. | Cytosol | [12] |

*, top-confident subcellular location.

Table A.2 The biomarkers and the surrogate transcriptome under pathological condition.

| Gene symbol | miRNA data availability | Tissue * | Condition * | Significant miRNA expression † | |
| --- | --- | --- | --- | --- | --- |
|  |  |  |  | Proportion (*n*/*N*, %) | Difference of regulation (*P*-value; miRNA OR, 95% CI) ‡ |
| ITGA5 | 26/27 (96.3%) | Placenta | PE | 8/26 (30.8) | *>*.05 |
|  |  |  | PE-FGR | 13/26 (50.0) | >.05 |
| P2RX7 | 38/39 (97.4%) | Placenta | PE | 8/38 (21.1) | >.05 |
|  |  |  | PE-FGR | 12/38 (31.6) | >.05 |
| IRF6 | 1/1 (100%) | Placenta | PE | 0/1 (0) |  |
|  |  |  | PE-FGR | 0/1 (0) |  |
| FANCI | 6/6 (100%) | Placenta | PE | 4/6 (67) | >.05 |
|  |  |  | PE-FGR | 3/6 (50) | <.001; (2/3; 67%):   1. hsa-let-7c-5p (MIRT051849) 0.03, 0.01 to 0.15 2. hsa-miR-192-5p (MIRT026117) 0.01, >0.00 to 0.11 |
| SELV | NA |  |  |  |  |
| TSEN15 | 3/3 (100%) | Placenta | PE | 2/3 (67) | >.05 |
|  |  |  | PE-FGR | 2/3 (67) | >.05 |
| TPX2 | 1/1 (100%) | Placenta | PE | 1/1 (100) | <.001; (1/1; 100%):   1. hsa-miR-193b-3p (MIRT041448) 7.2, 4.0 to 13.0 |
|  |  |  | PE-FGR | 0/1 (0) |  |
| WIPF3 | 11/11 (100%) | Placenta | PE | 3/11 (27.3) | >.05 |
|  |  |  | PE-FGR | 3/11 (27.3) | <.001; (2/3; 67%):   1. hsa-miR-1306-5p (MIRT643945) 12.0, 1.8 to 83.0 2. hsa-miR-6787-3p (MIRT643937) 8.4 × 10^3^, 7.8 to 9.1 × 10^6^ |
| ARID2 | 34/34 (100%) | Placenta | PE | 7/34 (20.6) | >.05 |
|  |  |  | PE-FGR | 10/34 (29.4) | >.05 |
| INSM1 | NA |  |  |  |  |
| TMEM38B | 2/2 (100%) | Placenta | PE | 1/2 (50) | <.001; (1/1; 100%):   1. hsa-miR-98-5p (MIRT027604) 0.18, 0.07 to 0.51 |
|  |  |  | PE-FGR | 1/2 (50) | <.001; (1/1; 100%):   1. hsa-miR-98-5p (MIRT027604) 0.05, 0.01 to 0.20 |
| ALS2CL | 15/16 (93.8%) | Placenta | PE | 3/15 (20) | >.05 |
|  |  |  | PE-FGR | 2/15 (13.3) | >0.05 |

*, query was conducted for all the genes in blood, cord blood CD8+ T cells, endometrium, amnion, myometrium, and placenta under conditions of preeclampsia, preeclampsia with intrauterine/fetal growth restriction, and preterm delivery; †, miRNA expression value in log_2_ reads per million (RPM); ‡, two-tailed permutation test in which either up- or down-regulation is significantly higher than that by chance across the genes; CI, confidence interval; FGR, fetal growth restriction; NA, not available; OR, odds ratio; PE, preeclampsia.

Table A.3 Interaction network connecting the biomarkers and the surrogate transcriptome.

| Inputs from the models * | | Predicted functional partners | | | | Network archived web page |
| --- | --- | --- | --- | --- | --- | --- |
| Gene symbol of the proteins | Tissue(s) | Step | Interactors † | | Connected inputs |  |
|  |  |  | 1^st^ shell | 2^nd^ shell |  |  |
| All proteins | All tissues | 1 | 0 | 0 | Incomplete | [13] |
|  |  | 9 | 50 | 50 | Incomplete, remaining SELV, INSM1, ALS2CL, TMEM38B |  |
| INSM1  ALS2CL  🡨  SELV  🡪  TMEM38B | Amnion (inner)  🡨  Cord (fetal) blood  🡪  Placenta (fetal side) | 1 | 0 | 0 | Incomplete | [14] |
|  |  | 8 | 50 | 20 | Complete |  |
| TPX2  WIPF3  ARID2  🡨  ITGA5  IRF6  🡪  PLOD2 ‡  TMEM38B | Decidua (maternal side)  🡨  Maternal blood  🡪  Placenta (fetal side) | 1 | 0 | 0 | Incomplete | [15] |
|  |  | 4 | 20 | 0 | Complete |  |

*, the best emulated biomarkers and the corresponding surrogate transcriptome in the PC-GBM; †, the maximum number of interactors were increased until all the inputs were connected, by default setting at the 1^st^ shell (0, 5, 10, 20, 50), then the maximum 50 interactors were maintained while increasing that at the 2^nd^ shell (0, 5, 10, 20, 50), if this the connection was still incomplete, then the remaining is considered to be the inputs for the next network; ‡, since no connection had been established between proteins in the maternal and fetal tissues, we added PLOD2 from the second network for the inputs to connect the proteins between decidua and placenta.

Table A.4 Subnetworks from inputs and the interactors in the connecting shortest paths.

| Proteins | Pair of tissues | | | | | | | | | | Network archived web page |
| --- | --- | --- | --- | --- | --- | --- | --- | --- | --- | --- | --- |
|  |  | Maternal blood | Cord (fetal) blood | | Decidua (maternal side) | | Fundus myometrium | Amnion (inner) | | |  |
|  | Maternal blood | (1) | (2) |  | (4) |  | (6) | (7) |  |  |  |
|  | Placenta (fetal side) |  |  | (3) |  | (5) |  |  | (8) |  |  |
|  | Cord (fetal) blood |  |  |  |  |  |  |  |  | (9) |  |
| ITGA5 | (input) | ✓ | ✓ |  | ✓ | ✓ | ✓ |  |  |  | [14-19] |
| P2RX7 | (input) | ✓ |  |  |  |  |  | ✓ |  |  | [16,17,20] |
| IRF6 | (input) | ✓ |  |  |  | ✓ |  | ✓ |  |  | [16,17,19,20] |
| GJA1 | (interactor) | ✓ |  |  |  |  |  |  |  |  | [16,17] |
| SERPINB5 | (interactor) | ✓ |  |  |  |  |  | ✓ |  |  | [16,17,20] |
| ITGB1 | (interactor) | ✓ | ✓ |  | ✓ | ✓ |  |  |  |  | [15-19] |
| FANCI | (input) |  | ✓ |  |  |  |  |  |  |  | [15,16] |
| TSEN15 | (input) |  | ✓ |  |  |  |  |  |  |  | [15,16] |
| TP53 | (interactor) |  | ✓ | ✓ |  |  |  |  |  | ✓ | [15,16,21,22] |
| ITGA2B | (interactor) |  | ✓ |  |  |  |  |  |  |  | [15,16] |
| TSEN54 | (interactor) |  | ✓ |  |  |  |  |  |  |  | [15,16] |
| TPX2 | (input) |  |  |  | ✓ | ✓ |  |  |  |  | [16,18,19] |
| WIPF3 | (input) |  |  |  | ✓ | ✓ |  |  |  |  | [16,18,19] |
| ARID2 | (input) |  |  |  | ✓ | ✓ | ✓ |  |  |  | [14,16,18,19] |
| TOP2A | (interactor) |  |  |  | ✓ | ✓ |  |  |  |  | [16,18,19] |
| ITGB3 | (interactor) |  |  |  | ✓ | ✓ |  |  |  |  | [16,18,19] |
| WASL | (interactor) |  |  |  | ✓ | ✓ |  |  |  |  | [16,18,19] |
| PBRM1 | (interactor) |  |  |  | ✓ | ✓ |  |  |  |  | [16,18,19] |
| PLK1 | (interactor) |  |  |  | ✓ | ✓ |  |  |  |  | [16,18,19] |
| TERT | (interactor) |  |  |  |  |  | ✓ |  |  |  | [14,16] |
| FN1 | (interactor) |  |  |  |  | ✓ | ✓ |  |  |  | [14,16,19] |
| INSM1 | (input) |  |  |  |  |  |  | ✓ | ✓ | ✓ | [16,20-22] |
| HDAC1 | (interactor) |  |  |  |  |  |  | ✓ |  | ✓ | [16,20,22] |
| TLR2 | (interactor) |  |  |  |  |  |  | ✓ |  |  | [16,20] |
| ALS2CL | (input) |  |  |  |  |  |  | ✓ | ✓ | ✓ | [16,20-22] |
| HMGB1 | (interactor) |  |  |  |  |  |  | ✓ |  |  | [16,20] |
| PIK3C3 | (interactor) |  |  |  |  |  |  | ✓ |  | ✓ | [16,20,22] |
| RAB5A | (interactor) |  |  |  |  |  |  | ✓ |  | ✓ | [16,20,22] |
| SELV | (input) |  |  | ✓ |  |  |  |  |  | ✓ | [16,21,22] |
| TMEM38B | (input) |  |  | ✓ |  | ✓ |  |  | ✓ |  | [16,19,21] |
| GPX4 | (interactor) |  |  | ✓ |  |  |  |  |  | ✓ | [16,21,22] |
| EZH2 | (interactor) |  |  | ✓ |  |  |  |  |  |  | [16,21] |
| PLOD2 | (interactor) |  |  | ✓ |  | ✓ |  |  | ✓ |  | [16,19,21] |
| KDM1A | (interactor) |  |  |  |  |  |  |  | ✓ |  | [16,21] |
| JMJD4 | (interactor) |  |  |  |  |  |  |  | ✓ |  | [16,21] |
| SMARCA4 | (interactor) |  |  |  |  | ✓ |  |  |  |  | [19] |
| KPNA2 | (interactor) |  |  |  |  | ✓ |  |  |  |  | [19] |

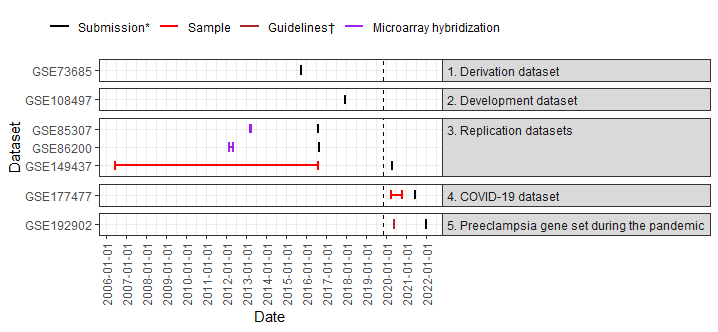

Figure A.1 Data period. Dashed line indicates the start date of COVID-19 pandemic worldwide (November 17, 2019). *, submission to gene expression omnibus (GEO); †, when collecting the data, preeclampsia in GSE192902 was defined by diagnostic criteria from the American College of Obstetrics and Gynecology (ACOG) guidelines that had been published on this date.

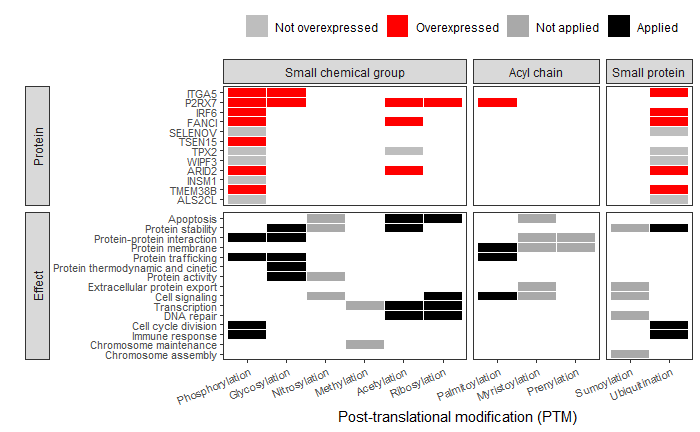

Figure A.2 The proteins and the PTMs with the biological effects. The PTM is classified into three group based on what the covalent attachment of. The PTMs with the effect is taken from Audagnotto and Dal Peraro (2017) except ribosylation. PTM, post-translational modification.
